# Nationwide Mpox Genomic Surveillance Reveals Clade Ib Introductions, APOBEC3-Driven Evolution, and Terminal Deletions

**DOI:** 10.64898/2026.07.15.26357894

**Authors:** Hayden N. Brochu, Qian Shi, Kuncheng Song, Qimin Zhang, Jason Munroe, Nathan J. Harris, Natalie Britt, Qiandong Zeng, Karan Kapuria, James Chappell, Brian M. Norvell, Lydia Peavy, Jonathan D. Williams, Ayla B. Harris, Jasmine Chaitram, Christina L. Hutson, Jiusheng Deng, Daisy McGrath, Deborah Boles, Suzanne E. Dale, Crystal M. Gigante, Lakshmanan K. Iyer

**Affiliations:** Labcorp, Burlington, NC 27215, USA; Laboratory Readiness and Informatics Branch, Division of Laboratory Systems, Office of Laboratory Systems and Response, Centers for Disease Control and Prevention, Atlanta, Georgia, USA; National Center for Emerging and Zoonotic Infectious Diseases, Centers for Disease Control and Prevention, Atlanta, Georgia, USA

**Keywords:** Mpox virus, whole genome sequencing, genomic surveillance, public health, clade IIb, clade Ib, APOBEC3, terminal deletion, probe-based sequencing, diagnostic integration

## Abstract

**Background:** The 2022-2023 global mpox outbreak highlighted the critical need for robust genomic surveillance capabilities to track mpox virus (MPXV) evolution and transmission dynamics.

**Methods:** Building upon our established SARS-CoV-2 sequencing infrastructure, we implemented a Molecular Loop® probe-based long-read sequencing approach using Pacific Biosciences Sequel II technology for comprehensive MPXV genomic surveillance across the United States (US). From August 2024 to June 2025, we generated 326 high-quality whole genome sequences from residual mpox-positive clinical specimens collected by Labcorp across all 10 US Department of Health and Human Services regions.

**Results:** Our analysis identified two samples containing clade Ib MPXV in January and June 2025 and captured shifting trends in clade IIb diversity, with 13 distinct lineages observed. We also identified multiple instances of large (∼1.6–17.6kb) deletions proximal to the inverted terminal repeats in clade IIb genomes. APOBEC3 mutation analysis indicated substantial evidence of human-to-human transmission among both clades. Further, we observed significantly higher APOBEC3-associated SNPs per kilobase (P<0.001) in clade IIb genomic variable regions relative to their central conserved region. Our assay exhibited strong reproducibility across biological replicates from individual patients and accuracy was confirmed via parallel sequencing of select specimens by US Centers for Disease Control and Prevention (CDC) using metagenomic sequencing. We also demonstrated via custom simulation that our assay discriminates all known MPXV clades and lineages, including those we have not observed in the US.

**Conclusions:** Our integrated nationwide surveillance system facilitates real-time genomic tracking of outbreak evolution, with demonstrated capacity across SARS-CoV-2 and MPXV, positioning this platform for rapid deployment during future pathogen emergence.

## Background

Mpox, a viral zoonotic disease caused by the mpox virus (MPXV; formally classified as belonging to *Orthopoxvirus monkeypox* by the International Committee on Taxonomy of Viruses), emerged as a significant public health concern in 2022 when a global outbreak predominantly involving clade IIb spread to over 100 countries, prompting the World Health Organization (WHO) to declare a Public Health Emergency of International Concern (PHEIC) in July 2022^1–4^. A consistent epidemiological feature of this clade IIb outbreak was the predominant transmission among those reporting male-to-male sexual contact^5^. While the initial PHEIC was lifted in May 2023, the subsequent emergence and international spread of clade Ib prompted WHO to declare a second PHEIC in August 2024^6^. The first detection of clade Ib MPXV in the United States was reported in California in November 2024 in a traveler returning from East Africa^7^. Together, these successive outbreaks underscore the ongoing global threat posed by MPXV and the critical need for robust genomic surveillance capable of tracking the emergence and spread of distinct viral clades and subclades^8,9^.

Genomic surveillance is critical for tracking pathogen evolution, identifying transmission chains, and detecting the introduction of novel variants^10^. These capabilities proved essential during the COVID-19 pandemic^11^ and remain vital for responding to emerging infectious diseases. MPXV, a double-stranded DNA virus with a ∼197 kilobase (kb) genome, presents sequencing challenges due to its large size, terminal inverted repeats, extensive tandem repeat regions, and genomic accordion-like expansions and contractions^12–15^. Successful MPXV whole genome sequencing (WGS) strategies thus far have used metagenomic- and amplicon-based approaches coupled with long read sequencing methods^16,17^. Moreover, commercial hybrid-capture panels, such as the Illumina® Viral Surveillance Panel and Twist Biosciences® Comprehensive Viral Research Panel, enhance MPXV metagenomic WGS by leveraging probe-design strategies exemplified by CATCH, a computational framework for optimizing hybrid-capture probe sets^18^. The accumulation of complete MPXV genome sequences throughout the clade IIb outbreak response resulted in more refined clade subtyping nomenclature^19^ and a new paradigm of APOBEC3 editing^20^ as a key driver of MPXV mutations during human-to-human transmission chains^13,21^.

In this study, we adapted our SARS-CoV-2 genetic surveillance infrastructure, which comprises diagnostic testing, sample sequestration, Molecular Loop® probe-based enrichment and long-read sequencing for high-throughput viral WGS^11^. We leveraged this adapted workflow to monitor ongoing clade IIb transmission and to detect potential clade Ib introductions, sequencing mpox-positive clinical specimens collected across all 10 United States Health and Human Services (US HHS) regions^22^. We further employed simulation methods, intra-assay reproducibility evaluations, and orthogonal (independent platform) comparisons with parallel collaborative US Centers for Disease Control and Prevention (CDC) sequencing efforts to validate our MPXV genetic surveillance apparatus. We investigated this genomic data to deliver key insights into the evolving MPXV epidemiological landscape in the US.

## Methods

### Mpox surveillance and WGS

Total nucleic acid was extracted from mpox-positive specimens identified using the US Food and Drug Administration (FDA)-cleared Labcorp® Mpox (*Orthopoxvirus*) PCR Test under Emergency Use Authorization (EUA) 230044^23^. These extracts were sequestered and consolidated via a Hamilton Microlab® STAR™ instrument. Amplification of the sample DNA was performed using a custom Molecular Loop® Mpox Virus (MPXV) capture kit^24^, which included 7,826 tiled Molecular Loop® Loopcap™ Molecular Inversion Probes (MIPs), covering ∼99.9% of the clade IIb MPXV genome with each base targeted by 26 MIPs on average. Each MIP averaged 20 base pairs (bp) in length, flanking a 675bp sequence between ligand and extension target sites. The resulting products were enriched and pooled at equal volumes.

Library preparation followed standard manufacturer protocols, including DNA damage repair, adapter ligation, enzymatic digestion to remove non-ligated products, and bead-based purification. Final libraries were sequenced on a Pacific Biosciences® Sequel II™ instrument^25^ with 15-hour movie runs.

### Sequence quality control and assignment of MPXV clades and lineages

Raw subreads were processed into circular consensus sequence (CCS) reads using the Pacific Biosciences^®^ SMRT LINK™ software v9.0 ccs program^26^. CCS reads were then demultiplexed using lima with the following parameters: “-min-score-lead −1”, “-min-score 80”, “-window-size-multi 1.1”, “-neighbors”. Since raw sequencing reads typically consist of ∼15 kb for the Pacific Biosciences^®^ Sequel II™^25^ and the sequencing product (including MIPs and barcodes) was ∼ 800 bp, each circular molecule underwent approximately 19 passes (15,000/800), yielding high confidence CCS reads. Molecular Loop® barcodes were trimmed using mimux v0.2.1 (PacBio HiFiViral Workflow) with the ‘-max-len 1500’ and ‘-min-score 0’ parameters. Final sample fastq files were generated by converting the resulting bam files using BamTools^27^.

Sequence fastq files were analyzed using a genome analysis pipeline tailored for MPXV implemented in the CLC Genomics Server version 9.1.1^28^. This consisted of processing sample-level fastq files using Minimap2 v2.17-r941^29^ to align reads to the MPXV clade IIb reference genome (NCBI GenBank reference NC_063383.1), generating a bam file of the alignment and VCF file containing the variants called using a custom variant caller in CLC. Consensus genomes were then generated using a reference-guided approach, VCFCons v8.5.0^30^, using the same reference genome as in the prior CLC analysis. First, samtools v1.3.1^31^ was used with ‘depth’ command and ‘-q 0 -Q 0’ parameters to calculate depth at all genomic positions. Unambiguous base calls required at least 4 CCS read coverage and an allele frequency consensus of at least 80%. If an alternate allele frequency was between 20% and 80%, the appropriate ambiguous IUPAC nucleotide was called. Nucleotides with coverage of less than 4 CCS reads were reported as ambiguous (N) in the consensus sequence.

Quality control (QC) was performed using two coverage calculations: 1) mean depth of coverage, which computes the average per-base depth of coverage across all genomic positions using samtools v1.3.1^31^ depth (described above) and 2) genome coverage, which computes the percentage of unambiguous genome base calls (i.e., not N) using the seqtk v1.2 ‘comp’ command^32^. MPXV genomes with at least 10X mean depth of coverage and 90% genome coverage were considered high-quality, passing QC and retained for further analysis in this study. High-quality MPXV genomes were further processed to determine clade and lineage assignment using Nextclade v3.14.2^33^. Clades were determined using the ‘All clades’ MPXV model (2025-04-25 release), which distinguishes MPXV sub clades (Ia, Ib, IIa and IIb) and further assigns clade IIb lineages.

### MPXV genome annotation

Before annotating MPXV genomes, terminal genomic Ns were trimmed off (i.e., removed) using the vadr v1.6.3^34^ fasta-trim-terminal-ambigs.pl script with ‘-minlen 0 -maxlen 1000000’ parameters. Next, liftoff v1.6.3^35^ was used to annotate genomes with RefSeq genes from their respective clade reference genome in NCBI GenBank (clade IIb: NC_063383.1, clade Ia: NC_003310.1). Gene annotation files (GFF files) were then processed in R v4.1.0^36^ using the tidyverse v.1.3.1^37^, seqinr v4.2.8^38^ and Biostrings v2.62.0^39^ packages. GFF files were parsed, then annotated regions were extracted from the corresponding MPXV genome sequence and translated to determine the presence of start and/or stop codons. If the translated sequence began with a start codon and ended with a stop codon without premature stop codons or frameshifts, then the annotation was considered ‘complete’ (otherwise ‘partial’). Gene annotations were also used to assess coverage across MPXV genomic regions, defined as the left variable region (V1), central conserved region (C), and right variable region (V2). The C region encompasses all RefSeq annotated genes from OPG056 to OPG151, V1 encompasses gene 5’ of OPG056, and V2 encompasses genes 3’ of OPG151^40^.

### Deletion detection and breakpoint refinement

Large deletions are challenging to detect using routine alignment and variant calling algorithms, including those employed in this study (see ‘Sequence quality control and assignment of MPXV clades and lineages’ Methods section). Therefore, a heuristic approach was employed to initially flag samples with high coverage (>10X mean depth, passing QC), yet significant evidence of read drop out concentrated in contiguous genomic locations. This was performed by evaluating the fractional depth of coverage at all genomic positions, calculated as the read depth at a position divided by the maximum read depth observed at any position. Positions with low fractional depth of coverage (<1%) were enumerated, and a minimum of 75 positions was required for further investigation (representing potential deletions ≥75 bp). Coverage profiles of flagged samples were analyzed in R v4.1.0^36^ using the Rsamtools v2.10.0^41^ and tidyverse v.1.3.1^37^ packages. Approximate deletion sizes and breakpoints were determined via manual inspection of read coverages in Integrative Genomics Viewer v2.16.2^42^.

IGV-annotated deletion coordinates were refined at base-pair resolution by an iterative alignment–pileup routine. For each putative deletion, a per-sample excised reference was built by splicing the annotated span out of the sample consensus, and a deletion-focused CCS read subset (reads within 5 kb of either breakpoint on the original consensus, plus all unmapped reads) was remapped to the excised reference with minimap2 v2.17-r941^29^ (-x map-pb). Local pileups spanning ±250 bp of the join (Rsamtools v2.10.0^41^ pileup) were classified as converged (deletion/insertion signal <10% and ≥5X spanning coverage), under-excised (deletion still present at the join), or over-excised (insertion at the join). Under-excised joins were refined by loading spanning alignments with GenomicAlignments v1.30.0 readGAlignments and extracting the modal reference span of the D-operator adjacent to the join across spanning reads (explodeCigarOps / explodeCigarOpLengths). Over-excised or low-coverage joins were routed to a *de novo* assembly path in which SPAdes v4.0.0^43^ contigs and/or subsampled reads were mapped to the original reference and breakpoints derived from the reference gap between primary and supplementary alignments of split reads, single-linkage clustered with ±200 bp tolerance. Refined breakpoints were accepted only when supported by ≥3 unique reads and the implied deletion length was within ±50% of the annotated span, suppressing spurious full-genome-spanning candidates from split alignments that switch between the near-identical inverted terminal repeats.

Breakpoints that could not be refined further were cross-checked against a full-sample *de novo* assembly (SPAdes v4.0.0^43^ --isolate) whose top scaffolds were mapped to both the original and excised references; a deletion was corroborated when the original-reference alignment recovered the deletion span (within ±25%) and the excised-reference alignment spanned the join with ≥50 bp of contiguous match on either side and no indel within ±20 bp. Two cases resolved on the basis of unfiltered contig alignments and per-base read coverage across the annotated join: 1) LC0000014 d1 was retained as assembly-implicit on the basis of a 158 kb scaffold whose primary M-run began 6 bp inside the right breakpoint, 2) LC0000014 d2 and LC0000184 d2 were retained on the coverage-cliff signature (per-base coverage dropping to zero immediately inside the annotated span and returning to sample-median coverage immediately after the annotated right endpoint). For the three F.1 same-donor samples (LC0000014, LC0000156, LC0000184), 1.5 kb of flanking sequence around each per-sample breakpoint was pairwise-aligned across the three donor consensuses (Biostrings v2.62.0^39^ pairwiseAlignment, type = "local") and each sample’s coordinates were projected onto the last shared homology position. Reconciled deletion lengths were identical at base-pair resolution with 100% flanking-sequence identity across all three samples. Final excised references were built by removing all accepted deletions from each sample consensus genome.

### Analysis of replicate sample concordance

Mpox genomic surveillance yielded two types of sample replicates: biological and technical. Biological replicates were multiple specimens collected from the same patient, whereas technical replicates were the same specimen sequenced multiple times. These sets of replicates (of size 2–4 samples) were compared based on their coverage metrics, annotation completeness, and clade/lineage assigned by the Nextclade^33^ ‘All clades’ model. In cases where replicate sets exceeded two samples, all samples were required to have the same clade and lineage to be considered concordant. Replicate sets were also assessed for concordance at mutation-level resolution using the Nextclade^33^ ‘All clades’ model mutation calls, separately for SNPs, insertions, and deletions. This analysis was constrained to complete, unambiguous (i.e., no N nucleotides) annotated genomic regions in all replicate set MPXV genomes. Any observed discrepancies were investigated by overlaying with tandem and inverted repeats detected in the genomes. Tandem repeats were determined using Tandem Repeats Finder v4.10.0^44^ with options ‘2 5 7 80 10 50 2000 -l 10 -d’. Inverted repeats were determined using Inverted Repeats Finder v3.09^45^ with options ‘2 3 5 80 10 40 500000 10000 -f -h -d -t4 74 -t5 493 -t7 10000’. Finally, replicate sets were de-deduplicated when investigating clade/lineage trends and when performing all phylogenetic and APOBEC3 mutation analyses. De-duplication was performed by choosing the genome with the highest depth of coverage as representative.

### CDC sequencing of selected mpox-positive samples and concordance analysis

Selected mpox-positive specimens were parallelly sequenced by CDC. Samples sent to CDC for parallel sequencing were selected based on practical criteria. Each sample required adequate residual specimen volume for repeat nucleic acid extraction. Chosen samples spanned a representative range of viral loads (Ct values). Selection was not based on lineage, geography, or any other genomic feature. Extracted DNA was used as input for the Illumina DNA Prep method according to the recommended protocol with ½ reagent volumes used throughout; samples were amplified using 5 or 11 PCR cycles depending on input concentration. Libraries were visualized using the Agilent Fragment Analyzer instrument and the HS NGS Fragment Kit (Agilent Technologies Inc., Santa Clara, CA). 48 to 64 libraries were pooled at approximately equal molarity and sequenced (200 pM final loading concentration) on an Illumina NovaSeq 6000 instrument using the 300-cycle SP sequencing components. MPXV genome sequences were assembled using PolkaPox v0.1-beta (https://github.com/CDCgov/polkapox), following the same procedures described in Gigante *et al*^46^.

CDC-sequenced genomes were evaluated for coverage, assigned a clade and/or lineage, and annotated using the same approaches as described in the ‘Sequence quality control and assignment of MPXV clades and lineages’ and ‘MPXV genome annotation’ Methods sections. The resulting MPXV genomes were analyzed for concordance with Labcorp-generated genomes using the same approach as in the ‘Analysis of replicate sample concordance’ Methods section.

### Time-calibrated phylogenetic analysis

Time-calibrated phylogenetic analysis of MPXV genome sequences was performed using augur v22.2.0^47^ and auspice.us v0.12.0 (Auspice 2.49.0) within the Nextstrain framework^48^. Since clade Ib is highly diverged from clade IIb and were insufficient in number for a standalone phylogeny, only clade IIb genomes were included in this analysis. The augur refine utility, which employs the TreeTime algorithm^49^, was used to perform time-calibrated phylogenetic analysis. This utilized molecular clock analysis, using coalescent optimization, marginal date inference, and a fixed clock rate of 6×10^-5^ substitutions/site/year (standard deviation 1×10^-5^) as recommended by Nextstrain MPXV^48^ and established for clade IIb by O’Toole *et al*^21^. Finally, this tree was rooted at the optimal position and exported to Auspice format for interactive visualization.

### Phylogenetic and APOBEC3 mutation analysis

APOBEC3 mutation analysis was performed for both clades, and phylogeny-aware APOBEC3 analysis also performed for clade IIb (insufficient samples for clade Ib). First, a reference sequence was selected for each clade. For clade IIb, we used the genome used by the Nextclade^33^ ‘All clades’ model (NCBI GenBank reference NC_063383.1), which was generated from a clade IIb sample collected in 2018. For clade Ib, the reference genome used by the Nextclade^33^ ‘clade I’ model was not used, as it was originally collected in 1979 (NCBI GenBank reference DQ011155.1). This genome pre-dates the recent clade Ib emergence^6^, so clade I genetic evolution would outweigh any recent APOBEC3-mediated activity from ongoing clade Ib outbreaks. Therefore, a more recently sequenced, high coverage genome from early in the Democratic Republic of Congo clade Ib outbreak was chosen (GISAID^50^ reference EPI_ISL_18886301: collected January 2024 from a hospitalized patient in South Kivu^51^).

All consensus MPXV genome sequences (from mpox-positive specimens collected between August 2024 and June 2025) were aligned to their respective clade reference genome using minimap2 v2.17-r941^29^ with default parameters. Single nucleotide polymorphisms (SNPs) were called using samtools v1.3.1 mpileup with parameters ‘-ugf’ followed by bcftools v1.3.1 with parameters ‘-vmo’^31^. A custom script in R v4.1.0^36^ was used with the seqinr v4.2.8^38^ and Biostrings v2.62.0^39^ packages to extract SNP APOBEC3 context (nucleotide upstream of SNP and full heptamer context). For combined assessment of potential APOBEC3-associated SNPs, G>A SNP contexts were reverse complemented to match the same strand as T>C SNPs. SNP frequencies were compared across the MPXV genomic regions (V1, C, V2) by standardizing the total SNPs in a region using the length of the region.

APOBEC3 analysis was also performed using a phylogeny-aware approach, Squirrel v1.1.16^52^, which places SNPs in the phylogeny. Only clade IIb genomes were used in this analysis, and this was confirmed using the ‘-clade split’ argument, which separates clade I and II sequences (no clade I detected). Additionally, the ‘-run-apobec3-phylo’ parameter was used for identifying APOBEC3-associated mutation signatures of sustained human-to-human transmission. Further, the ‘-seq-qc’ QC parameter was used to flag potentially problematic sequences and sites. The resulting phylogenetic tree was annotated using the clade IIb lineage assignments.

### Statistical analysis

Statistical comparisons of mutation densities across MPXV genomic regions (V1, C, V2) were performed using Wilcoxon signed-rank tests, which evaluated paired differences in SNP densities per kb within individual samples. Multiple hypothesis testing correction was applied using the Benjamini-Hochberg procedure to control the false discovery rate. All statistical analyses were performed in R v4.1.0^36^, and *P* values < 0.05 were considered statistically significant after correction for multiple comparisons.

### Labcorp MPXV WGS simulation design

The Labcorp MPXV WGS assay simulator was designed to generate CCS reads from a ground truth MPXV genome sequence (see ‘Simulation data collection and testing’ Methods section). Such input genomes should be complete with minimal sequence ambiguity (i.e., few Ns). The simulation logic follows the assay design by incorporating a probe selection routine based on their alignment identity with the ground truth genome. The following procedure is employed to yield a CCS fastq file for an input MPXV ground truth genome. The full procedure utilized python v3.12^53^ and at minimum required biopython ≥v1.84^54^, pandas ≥v2.2.3^55^ and numpy ≥v2.1.2^56^:

1. **MIP alignment to the ground truth genome.** This utilized VSEARCH v2.17.1^57^ with ‘usearch_global’ command and ‘-id 0.5 -strand both -maxgaps 0 -minwordmatches 0 - maxaccepts 0 -userfields query+target+id+alnlen+qstrand+mism+opens+qrow+trow+caln’ options. Since MPXV genomes exceed the maximum reference length allowed by VSEARCH (∼190kb versus 50kb maximum), genomes were preprocessed before alignment to chunk them. This procedure used a sliding window approach to generate up to 50kb chunks with 50bp overlaps (e.g., chunk 1: positions 1–50,000; chunk 2: positions 49,051–100,050; etc.), with original genomic coordinates maintained for subsequent MIP fill-in extraction.
2. **MIP alignment processing and selection.** MIP alignments were processed to extract valid ligand-extension MIP arms by requiring valid strand alignment (ligand on positive strand and extension on negative strand or vice versa). Also, each MIP arm was required to have at most three mismatches. Since MIPs with perfect or near-perfect matches to the target sequence are more likely to successfully capture and amplify target regions, an inverse relationship with MIP arm mismatch counts was used for probabilistic selection of probes—*1/(n_lig+n_ext+1)*—where *n_lig* and *n_ext* signify the number of mismatches in the ligand and extension arms, respectively. The sum of these ratios was used to scale probe selection probabilities. The selection routine was applied to randomly draw from the valid MIP arm pairs, where the total number of selections was calculated using user-supplied target depth of coverage calculated as target depth x ground truth genome length divided by the average fill-in sequence length (675bp, see ‘Mpox surveillance and WGS’ Methods section).
3. **CCS fastq file generation.** For each MIP arm pair with at least one selection, the fill-in was extracted from the ground truth genome sequence using the alignment coordinates of each arm. An empirical approach was employed for assigning read qualities by randomly selecting read quality strings from a read archive compiled from sequencing runs in this study. If extracted fill-ins exceeded the length of the drawn quality string, then additional quality string(s) were drawn and concatenated until the quality string covered the fill-in sequence length. Once completed, the quality string was trimmed to match the sequence length. Finally, the fill-in sequence was mutated probabilistically using the quality string Phred scores for each base, where the quality encoding followed standard Phred+33 format: Q = −10 x log_10_(P_error). If an error was selected, then a random alternative nucleotide was chosen. The resulting mutated sequences and matched quality strings were compiled as a fastq file.

### Simulation data collection and testing

The NCBI Virus database^58^ was queried using the NCBI datasets v18.0.2 command-line tool with the command and parameters ‘download virus genome taxon 10244’ on June 5^th^, 2025 to retrieve all relevant MPXV clade and lineage genomic data for simulation testing. Of these 10,417 downloaded genomes, we selected a randomized subset of ≤20 genomes per country, yielding 670 for further evaluation. Next, these genomes were assigned clades and lineages using Nextclade v3.14.2^33^ and the ‘All clades’ MPXV model (2025-04-25 release), as done for the genomes sequenced in this study. The final set of genomes to be simulated were selected by prioritizing high coverage genomes with length ≥185 kb for each unique clade and lineage. For clades Ia, Ib, and IIa, up to five genomes were selected. For clade IIb, up to two genomes were selected for each lineage. This process yielded a final set of four clade Ia, five clade Ib, five clade IIa, and 65 clade IIb genomes across 35 lineages for simulation analysis (**Supplementary Data**).

All 79 MPXV genomes were simulated using four target sequencing depths (5X,10X, 20X, 30X). For each unique combination of genome and depth, three replicates were generated using different random seeds (1–99) for the MIP pair selection and read error introduction procedures. So, in total 240 genomes were simulated at each sequencing depth, yielding a total of 960 simulated CCS fastq files. All fastq files were processed through the steps described in the ‘Sequence quality control and assignment of MPXV clades and lineages’ Methods section, beginning after the probe trimming step (probe arms not included in the simulated fastq). For each sample, the simulated mean depth of coverage was compared to the target depth, and the clade/lineage assignment was compared to that of the ground truth genome. Full simulation results with coverage data, Nextclade^33^ clade/lineage assignments, and ground truth genome information are provided in the **Supplementary Data**.

## Results

### Nationwide sample collection spanning all US HHS regions

Adapting our SARS-CoV-2 surveillance apparatus^11^ for MPXV, we successfully sequenced 326 residual mpox-positive clinical specimens collected from 288 unique patients with demographic data available between August 2024 and June 2025, representing all 10 US HHS regions (**Figure 1a**). All sequenced MPXV genomes in this study were required to pass stringent QC, requiring 10X mean depth of coverage and >90% genome coverage (Methods). The geographic distribution generally followed our SARS-CoV-2 surveillance trends with higher sample counts near ports of entry in high population density regions, such as New York (n=56; 19.4% of 288) and California (n=71; 24.7% of 288) in HHS regions 2 and 9, respectively (**Figure 1a**). Other notable surveilled regions spanned the remaining eastern seaboard, consisting of HHS regions 4 (n=41) and 3 (n=27), and HHS region 6 comprising Texas and neighboring states (n=38) (**Figure 1a**). Patient demographics showed predominant transmission among men (n=277, 96.2% of 288) with patients most commonly in their 30s (n=118, 41% of 288) (**Figure 1b**), consistent with previously reported mpox epidemiology predominantly affecting men who have sex with men during the 2022-2023 US outbreak^5^. Specimen types were primarily labelled as viral transport media (n=231, 80.2% of 288) and swabs (n=26, 9% of 288), with cases seldomly providing more precise anatomical sources, such as penis, back, and rectum (**Figure 1c**). Sample cycle threshold (Ct) values from the VAC1 target of the Labcorp Mpox PCR assay ranged from 16 to 34 (median=23.4; **Figure S1**), demonstrating successful genome recovery across a range of viral loads.

**Figure 1.**
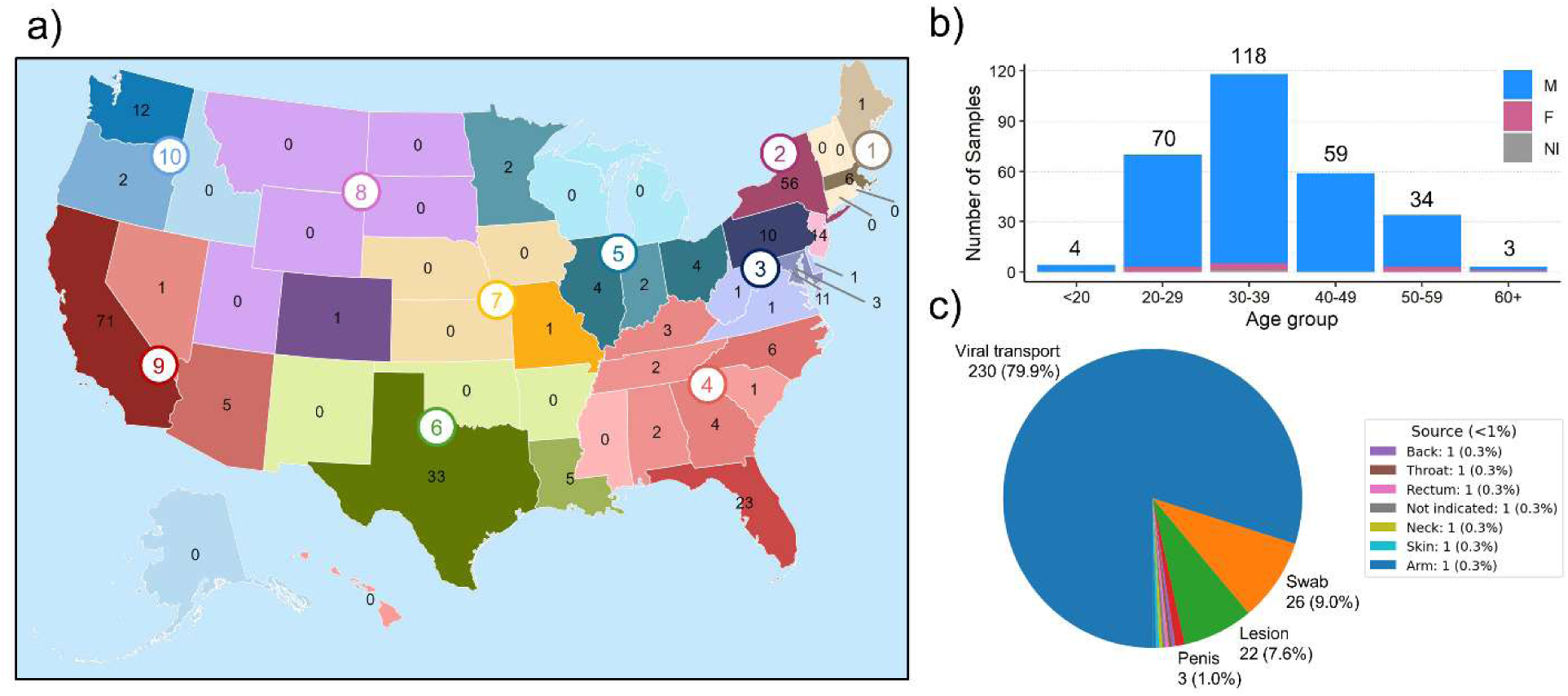
Surveillance and demographic overview of sequenced mpox-positive cases. A total of 326 specimens were sequenced, representing 291 unique patients. Of these, 288 had available metadata and are visualized here. **(a)** Geographic distribution of sequenced samples across the US. States are shaded according to sample count (darker shades indicate higher counts), with color coding by US HHS region (1-10). **(b)** Patient age distribution with stacked bar plots representing the number of samples per age group separated by sex: blue for male (M), pink for female (F), and gray for not indicated (NI). Total numbers of each age group are shown above the stacked bars. **(c)** Composition of sample types and origins. Pie chart displays the proportional breakdown of specimen types included in the final dataset with both number and percentage of total shown.

### Genomic surveillance shows dynamic lineage shifts and detection of clade Ib

We then analyzed all de-duplicated MPXV genomes from 288 unique patients to assess temporal and phylogenetic trends (**Figure 2**). Most sequences were assigned to clade IIb lineages (**Figure 2**; 286, 99% of 288), consistent with the dominant circulating clade since the 2022 outbreak. The other two samples were both identified as clade Ib (marked with asterisks in **Figure 2c**), representing critical surveillance detections with one case each in January 2025 (deposited in collaboration with CDC; NCBI GenBank PV448277.1) and June 2025 that we swiftly reported to CDC for immediate public health response. These cases were determined to be isolated U.S. introductions among recent travelers to countries with ongoing clade Ib outbreaks.

**Figure 2.**
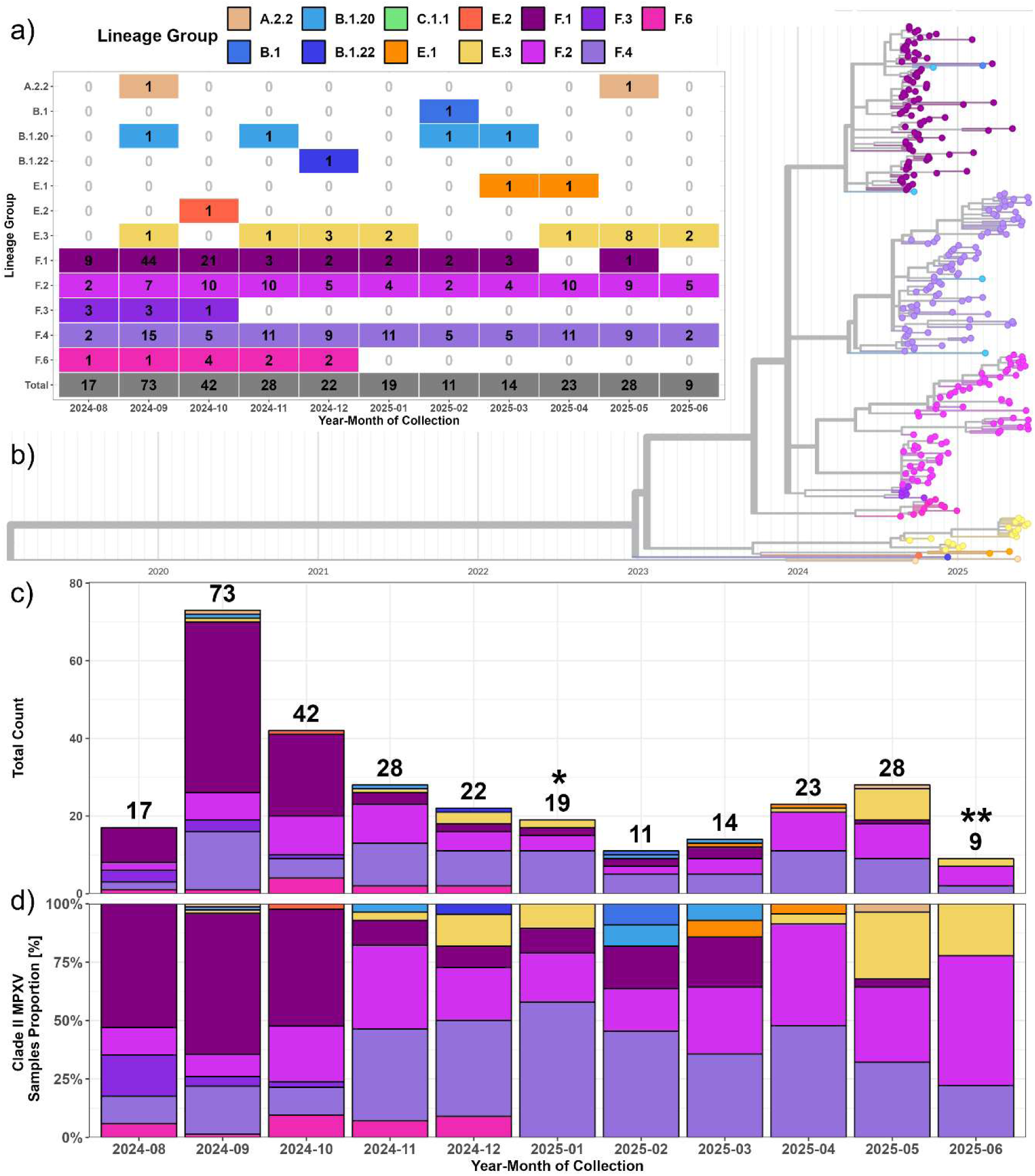
MPXV phylogenetic and temporal trends in the US between August 2024 and June 2025. The 13 unique clade IIb lineages detected are colored consistently across each panel. **(a)** Numbers of lineage detections across each month of surveillance. **(b)** Time-calibrated phylogenetic tree generated with augur^47^ with a clock-rate set to 6×10^-5^ mutations per site and NCBI GenBank reference NC_063383.1 as the root (collected August 2018). **(c-d)** Stacked bar plots of monthly lineage counts (c) and proportions (d) with clade Ib detections indicated by asterisks (*One case in New York, **One case in Massachusetts). Total monthly numbers are shown above the bars in (c).

Temporal analysis of clade IIb samples revealed dynamic lineage composition shifts over the study period (**Figure 2**). Lineage F.1 predominated from August through October 2024, while F.2 and F.4 became the majority lineages from November 2024 onwards. The dominance of the F.1 lineage coincided with a surge in surveillance volume from September through October 2024 (**Figure 2**), likely reflecting increased case incidence rather than sampling intensity, as our specimen collection remained consistent with diagnostic testing volumes. Notably, lineage E.3 showed increased prevalence in the latest surveillance months, particularly in May and June 2025 (**Figure 2**). Lineage A variants were rare, with only A.2.2 detected once in both September 2024 and May 2025 (**Figure 2**). This was expected as lineage A variants are not known to circulate in the US. The time-calibrated phylogeny confirmed that Nextclade-assigned^33^ lineages formed well-supported monophyletic clusters with temporal structure (**Figure 2b**), validating the lineage classification nomenclature and demonstrating ongoing viral evolution. Additionally, our time-calibrated phylogeny placed the divergence between F.1, F.3, and F.4 groups in early 2024, indicating that our surveillance network captured ongoing evolution within these distinct MPXV variants well after their initial emergence.

### High-coverage MPXV genomes enable detection of rare large terminal deletions

Sequencing coverage analysis across all samples (including replicates, n=326) demonstrated robust genome recovery (minimum 10X mean depth of coverage, 90% genome coverage) with near-100% complete genomes at 100X depth (**Figure 3a**). Median coverage metrics were remarkably high (261X depth, 99.8% coverage) with the detected clade and lineage having minimal impact on genome annotation completeness (**Figure 3b**). The relationship between Ct values and our coverage metrics showed the expected inverse correlation, with depth of coverage gradually dropping as Ct values exceeded ∼22 (**Figure S1**). However, we were still able to obtain high coverage (>99%) genomes even for samples with Ct values approaching 30 (**Figure S1**).

**Figure 3.**
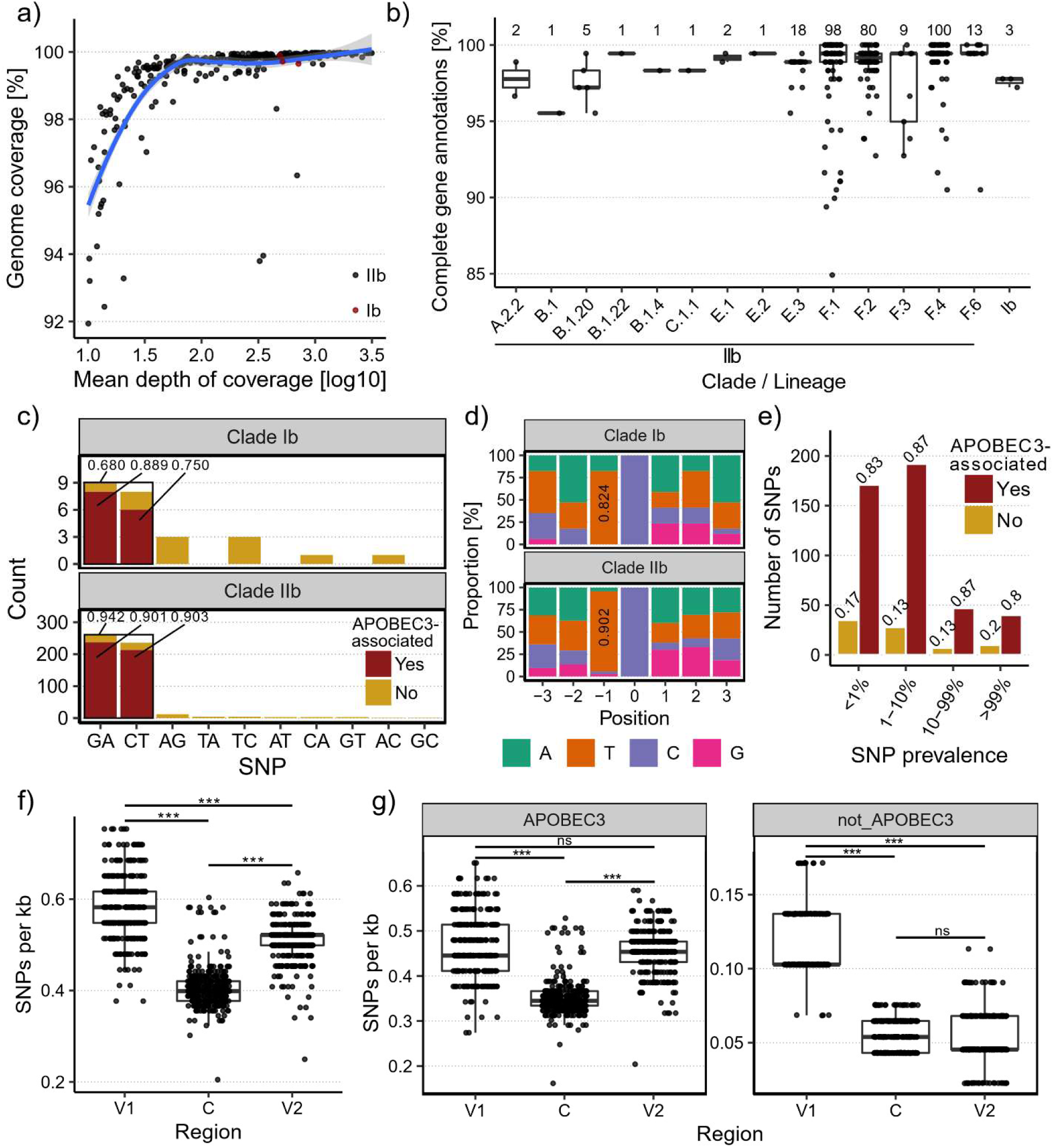
Genome coverage, annotation, and APOBEC3-associated mutation analysis of 326 high-quality MPXV genomes. **(a)** Scatterplot of mean depth of coverage [log10, x-axis] versus Genome coverage [%, y-axis] for each genome, colored by clade (black: IIb, red: Ib). The blue line indicates a loess curve with 95% confidence interval as gray fill. **(b)** Boxplots indicating the genes annotated [%] for each genome, stratified by clade/lineage with numbers of genomes in each boxplot shown above. **(c)** Number of SNPs detected (e.g., ‘GA’ indicates a G>A mutation), stratified by clade (top: Ib, bottom IIb). G>A and C>T mutations are further separated based on whether they were APOBEC3-associated (red, proportion shown) or not (yellow), i.e., dinucleotide mutations GA>AA and TC>TT, respectively. The proportion of SNPs that were G>A or C>T is also shown. **(d)** Stacked bar plot showing heptamers of G>A and C>T SNPs, with G>A reverse complemented to the negative strand to coerce the same context with ‘C’ at position 0. **(e)** SNP frequencies stratified by prevalence and whether they were APOBEC3-associated (red) or not (yellow). Proportions are shown for each bar within each prevalence range. **(f-g)** SNPs per kb across conserved (C) and variable regions (V1, V2) overall (f) and stratified by APOBEC3 association (g). The C region encompasses all RefSeq annotated genes from OPG056 to OPG151, V1 encompasses gene 5’ of OPG056, and V2 encompasses genes 3’ of OPG151. \*\*\**P*<0.001; ns=not significant.

A few samples exhibited coverage anomalies upon visual inspection of genome coverage as it relates to mean depth of coverage (**Figure 3a**). We further investigated these by quantifying the number of genomic positions with significant drops in read coverage. By enumerating genomic positions with a fractional coverage <1% (i.e., coverage as a fraction of maximum coverage across all genomic positions), we identified 10 samples with clear evidence of dropout (**Figure S2a**). Further evaluation indicated that there were large (∼1.6–17.6 kb) deletions near or within the inverted terminal repeats, highly distinct from the more even coverage across the rest of these MPXV genomes (**Figure S2b**). All putative deletions were then manually verified by visual inspection of read alignment patterns in IGV^42^ (**Figure S3**) and subsequently refined at base-pair resolution by an iterative alignment-pileup routine on per-sample excised references (Methods; **Figure S4a**; **Table 1**). Three F.1 samples all collected from the same patient contained two deletions: a ∼10.4 kb deletion encompassing OPG015 within the left inverted terminal repeat (ITR1) and six adjacent genes, and a ∼1.8 kb deletion encompassing OPG015 within the right terminal inverted repeat (ITR2) and part of OPG016 (**Figure S3a-b**; samples 4, 5, 8; **Table 1**). Reconciling the three same-donor sample breakpoints to shared flanking homology yielded identical deletion lengths (10,410 bp and 1,775 bp) at base-pair resolution across all three, with 100% flanking-sequence identity, confirming that they represent the same two biological deletion events (**Figure S4d**). One F.4 sample contained a similar ITR1-proximal ∼7.6 kb deletion affecting six genes, though with different start and end points (**Figure S3a**; sample 9; **Table 1**). Four samples (three F.4, one F.2) contained an ITR2-proximal ∼17.5 kb deletion affecting 7 genes (**Figure S3c**; samples 1, 3, 7, 10; **Table 1**). Finally, two F.4 samples had a pair of deletions: one ∼8.2 kb near ITR2 and one ∼1.7 kb near ITR1 (**Figure S3c-d**; samples 2, 6; **Table 1**). Overall, these large deletions were rare with detection in only 2.7% (8 of 291) of unique mpox-positive cases and exhibited similar characteristics to those recently described by Gigante *et al*^46^ for B.1 MPXV deletions during the 2022 outbreak in the US, including deletion locations near ITRs and affected gene sets.

**Table 1.** Refined breakpoints and evidence tier for the 14 large terminal deletions detected across 10 samples with read coverage dropout. Deletion coordinates (del_left, del_right) were obtained by iteratively refining the initial IGV-based breakpoint calls (**Figure S3**) against per-sample excised references (Methods, "Deletion breakpoint refinement"; **Figure S4a**). Deletion coordinates are 1-based, inclusive, on each sample’s own consensus genome and are shown in ascending order (del_left). ITR side indicates whether the deletion lies within/adjacent to the left (ITR1) or right (ITR2) inverted terminal repeat, as identified by proximity to positions 1–1,921 or 195,873–197,209 on the reference Clade IIb genome. Approximate deletion sizes are rounded to the nearest 100 bp, and a reconciled base-pair length is available for the three same-donor samples (†). Exact reconciled lengths are in parentheses (see **Figure S4d**). Evidence tier summarizes how the refined breakpoints were corroborated (Methods, "Deletion breakpoint refinement").

| Sample (label) | Deletion | ITR side | del_left | del_right | Approx. size | Evidence Tier |
| --- | --- | --- | --- | --- | --- | --- |
| LC0000161 (1) | d1 (1/1) | ITR2 | 173,201 | 190,730 | ~17.5 kb | ITR-caveat |
| LC0000158 (2) | d1 (1/1) | ITR2 | 173,162 | 190,724 | ~17.6 kb | ITR-caveat |
| LC0000276 (3) | d1 (1/1) | ITR2 | 173,169 | 190,733 | ~17.6 kb | ITR-caveat |
| LC0000156 (4) † | d1 (1/2) | ITR1 | 4,616 | 14,664 | ~10.0 kb (10,410 bp reconciled) | Pileup-converged |
| LC0000156 (4) † | d2 (2/2) | ITR2 | 190,828 | 192,456 | ~1.6 kb (1,775 bp reconciled) | Pileup-converged (1 bp shift) |
| LC0000014 (5) † | d1 (1/2) | ITR1 | 4,123 | 14,527 | ~10.4 kb (10,410 bp reconciled) | Assembly-implicit |
| LC0000014 (5) † | d2 (2/2) | ITR2 | 190,543 | 192,276 | ~1.7 kb (1,775 bp reconciled) | Coverage-cliff |
| LC0000235 (6) | d1 (1/1) | ITR1 | 14,272 | 15,886 | ~1.6 kb | Pileup-converged |
| LC0000011 (7) | d1 (1/2) | ITR1 | 14,296 | 15,962 | ~1.7 kb | Pileup-converged |
| LC0000011 (7) | d2 (2/2) | ITR2 | 182,579 | 190,721 | ~8.1 kb | Pileup-converged |
| LC0000184 (8) † | d1 (1/2) | ITR1 | 4,444 | 14,548 | ~10.1 kb (10,410 bp reconciled) | Pileup-converged |
| LC0000184 (8) † | d2 (2/2) | ITR2 | 190,643 | 192,417 | ~1.8 kb (1,775 bp reconciled) | Coverage-cliff |
| LC0000104 (9) | d1 (1/1) | ITR1 | 6,382 | 13,987 | ~7.6 kb | Pileup-converged |
| LC0000260 (10) | d1 (1/1) | ITR2 | 182,555 | 190,715 | ~8.2 kb | Pileup-converged |

† Same-donor biological replicates (LC0000014, LC0000156 and LC000018).

### Extensive APOBEC3-mediated mutagenesis drives MPXV clade IIb evolution and is detected among clade Ib introductions

Because we obtained high coverage and complete annotations across our MPXV genomes (**Figure 3a-b**), we were motivated to investigate genome-wide patterns of APOBEC3-associated SNPs, which are a product of sustained human-to-human transmission^21^. We analyzed APOBEC3 mutation patterns within MPXV coding regions across both clades, using the clade IIb reference genome used by Nextclade^33^ (NCBI GenBank reference NC_063383.1) and a high coverage clade Ib genome from the recent Democratic Republic of Congo clade Ib outbreak (GISAID^50^ accession EPI_ISL_18886301) (**Figure 3c-d**; Methods). These reference genomes were selected to ensure APOBEC3 signatures reflected contemporary human-to-human transmission rather than historical zoonotic evolution. As expected, we observed substantial evidence of APOBEC3-associated activity among G>A (90.1%) and C>T (90.3%) SNPs in clade IIb, with G>A and C>T SNPs combining for 94.2% of all clade IIb SNPs detected (**Figure 3c**). We also observed APOBEC3-associated SNPs prevalent among the two clade Ib genomes we sequenced, with 88.9% of G>A and 75% of C>T SNPs exhibiting the characteristics of APOBEC3 deamination (**Figure 3c**). Reverse complementing G>A SNPs to the negative strand, combining with C>T SNPs, and analyzing their full heptamer genomic contexts yielded overall measures of 90.2% and 80.4% APOBEC3 activity for clades IIb and Ib, respectively (**Figure 3d**).

We also assessed the temporal, phylogenetic and genome-wide characteristics of clade IIb APOBEC3 activity (**Figure 3e-g**). We first binned SNPs by prevalence to evaluate how APOBEC3 signatures have persisted throughout our surveillance, finding strong (>80%) activity across the full spectrum analyzed (**Figure 3e**; <1%, 1-10%, 10-99%, and >99% prevalence). As expected for an actively evolving viral population, the majority of unique SNPs were low-frequency variants detected in <1% of genomes (38%, 171 of 450 SNPs) or 1-10% of genomes (42.7%, 192 of 450 SNPs), with strong APOBEC3 signatures persisting across all frequency bins (**Figure 3e**). Phylogenetic analysis showed APOBEC3 activity spanning all clade IIb lineages, highlighting this process as a primary driver of ongoing clade IIb evolution and lineage diversification in the US (**Figure S5**).

We then analyzed the impact of this substantial mutation burden on the conserved core of the MPXV genome (C) compared to the left-(V1) and right-flanking (V2) variable regions (**Figure 3f-g**, Methods). A significantly higher density of overall SNPs was observed per kb in both V1 (median ∼0.6) and V2 (median ∼0.5) compared to C (median ∼0.4) (**Figure 3f**, Wilcoxon signed-rank tests, both *P*<0.001). Notably, V1 exhibited significantly higher SNPs per kb than V2 (median ∼0.6 vs ∼0.5; **Figure 3f**, Wilcoxon signed-rank test, *P*<0.001), suggesting potential asymmetry in replication dynamics or repair mechanisms between the terminal genomic regions. This differential mutation density was not attributable to coverage bias, as pairwise correlations of mean depth across genomic regions within sample were extremely strong (**Figure S6**, all Spearman ρ > 0.99). Stratification of SNPs by APOBEC3 association confirmed that APOBEC3 activity was significantly lower in the conserved core relative to variable regions (**Figure 3g**, Wilcoxon signed-rank tests, both *P*<0.001). The V1 region also had a significantly higher density of SNPs not related to APOBEC3 compared to both C and V2, while C and V2 exhibited similar rates (**Figure 3g**, Wilcoxon signed-rank tests, both *P*<0.001). Together, these results indicate that APOBEC3 activity is accelerating genome-wide clade IIb evolution with less tolerance in the conserved core of the genome.

To evaluate whether these regional differences were driven by the ITRs, we repeated the analysis after masking ITR sequences in V1 and V2. The overall differences in regional mutation density remained significant, though the median difference between V1 (0.493) and V2 (0.433) was smaller (**Figure S7a**, Wilcoxon signed-rank tests, all *P*<0.001). However, for APOBEC3-associated mutations, V1 and C no longer differed significantly (*P*=0.247), while V2 was significantly higher than both V1 and C (both *P*<0.001), indicating that APOBEC3-driven mutagenesis in V1 is partly influenced by ITR content (**Figure S7b**, Wilcoxon signed-rank tests). Together, these results indicate that APOBEC3 activity is accelerating genome-wide clade IIb evolution with reduced tolerance in the conserved core of the genome.

### Biological and technical replicates exhibit strong reproducibility

Our MPXV surveillance often involved multiple clinical specimens collected from individual patients, providing us with an opportunity to evaluate the reproducibility of our MPXV WGS assay. Of the 326 high-quality MPXV genomes in this study, 53 (16.3%) were replicates across 23 patients (‘biological replicates’; 2–4 per patient), and eight (2.5%) were patient samples sequenced in duplicate (‘technical replicates’) (**Table 2**). Biological replicates included specimens from the same anatomical site when available, as well as specimens from different sites on the same patient, providing insights into both technical reproducibility and potential intra-host genomic variation. Lastly, all replicates were from Clade IIb mpox-positive cases.

**Table 2.** MPXV WGS assay reproducibility across biological and technical replicates. Table is split first by replicate type (biological: same patient, different specimen; technical: same specimen, different sample). Replicates have 2–4 samples, with each sample result semicolon-delimited. Results are shown for Clade IIb lineages assigned by Nextclade^33^, mean depth of coverage, genome coverage [%], and number of complete gene annotations.

| Replicate |  |  | Nextclade lineage | Mean depth of coverage | Genome coverage [%] | # Complete annotations |
| --- | --- | --- | --- | --- | --- | --- |
| Type | ID | # Samples |  |  |  |  |
| Biological | bio1 | 3 | F.1;F.1;<br>F.1 | 49.2;25.4;<br>12.4 | 99.6%;98.7%;<br>95.2% | 177;175;<br>161 |
|  | bio2 | 2 | F.4;F.4 | 647.4;204.6 | 99.9%;99.8% | 178;178 |
|  | bio3 | 2 | F.4;F.4 | 60.5;27.1 | 99.4%;99.3% | 177;177 |
|  | bio4 | 2 | F.4;F.4 | 885.3;21.2 | 99.9%;97.8% | 178;175 |
|  | bio5 | 3 | F.1;F.1;<br>F.1 | 13.9;349;<br>20.6 | 92.4%;94%;<br>93.3% | 164;169;<br>167 |
|  | bio6 | 2 | F.2;F.2 | 56.1;36.5 | 98.5%;98.4% | 174;173 |
|  | bio7 | 2 | F.4;F.4 | 714.1;54.4 | 99.7%;99.6% | 178;178 |
|  | bio8 | 2 | F.1;F.1 | 412.3;687.8 | 99.9%;100% | 178;179 |
|  | bio9 | 2 | F.2;F.2 | 204.5;20.2 | 99.8%;99% | 178;175 |
|  | bio10 | 4 | F.4;F.4;<br>F.4;F.4 | 134.8;70.3;<br>39.4;27.9 | 99.7%;99.8%;<br>98.5%;99.7% | 178;178;<br>168;178 |
|  | bio11 | 4 | F.1;F.1;<br>F.1;F.1 | 503.7;361.4;<br>121.8;302.5 | 99.9%;99.9%;<br>99.8%;100% | 179;178;<br>178;179 |
|  | bio12 | 2 | F.4;F.4 | 70.7;75.6 | 99.7%;99.9% | 178;178 |
|  | bio13 | 2 | F.4;F.4 | 316;10.4 | 99.8%;93.2% | 178;162 |
|  | **bio14** | **2** | F.4;F.4 | 919.1;25.7 | 99.9%;99.4% | 179;177 |
|  | **bio15** | **2** | F.4;F.4 | 229.3;140.7 | 99.9%;99.9% | 179;179 |
|  | **bio16** | **3** | F.6;F.6;<br>F.6 | 833.2;566.5;<br>430.1 | 99.8%;99.8%;<br>99.8% | 178;178;<br>178 |
|  | **bio17** | **2** | F.2;F.2 | 145.3;95.3 | 99.7%;99.4% | 177;176 |
|  | **bio18** | **2** | F.4;F.4 | 806.3;678.7 | 99.9%;99.9% | 178;179 |
|  | **bio19** | **2** | F.4;F.4 | 1138.1;566.2 | 100%;99.8% | 179;178 |
|  | **bio20** | **2** | F.2;F.2 | 1711.1;626 | 100%;99.8% | 179;178 |
|  | **bio21** | **2** | F.6;F.6 | 3014.4;115.5 | 100%;99.7% | 179;178 |
|  | **bio22** | **2** | F.2;F.2 | 1291.9;494.6 | 99.9%;99.7% | 179;178 |
|  | **bio23** | **2** | F.4;F.4 | 89.7;66.1 | 99.2%;99.6% | 169;178 |
| **Technical** | **tech1** | **2** | F.1;F.1 | 1501.9;371.1 | 100%;100% | 179;179 |
|  | **tech2** | **2** | F.4;F.4 | 562.3;418.4 | 99.9%;99.9% | 178;178 |
|  | **tech3** | **2** | F.2;F.2 | 1828.6;704.7 | 100%;99.9% | 179;178 |
|  | **tech4** | **2** | F.2;F.2 | 184.8;89.6 | 99.8%;99.7% | 178;178 |

We observed perfect lineage concordance across both biological and technical replicates, despite some differences in coverage metrics and genome annotation completeness (**Table 2**). A more granular examination of mutation calls showed 100% SNP concordance (median) with a 98.5%–100% interquartile range (IQR) for biological replicates and 100% concordance for all technical replicates (**Table S1**). Deletions, though rarely detected in genic regions, were perfectly concordant in all biological and technical replicates (**Table S1**). Genic insertions were only observed within a single tandem repeat region (ATC repeat at positions 136,513–136,569 in reference NC_063383.1) detected across 6 biological replicates, with only half of replicates showing concordance for insertions calls, indicating some limitations in sequencing accuracy in such low complexity regions. Taken together, these results show that our MPXV WGS assay generated high-fidelity genomes with strong reproducibility, and they also indicate that MPXV genetic variation is minimal across sample collections from the same patient. This is consistent with prior intra-host studies of clade IIb during the 2022 outbreak in New York City^59^, Melbourne^60^, and across multiple body tissues over time^61^. While those studies identified low-frequency intra-host minority variants by deep metagenomic sequencing, our consensus-level analysis indicates that any such variation rarely reaches the consensus threshold, supporting the use of a single specimen per patient for lineage assignment and phylogenetic placement.

### Labcorp MPXV WGS accuracy confirmed via sequencing at CDC

As part of our efforts to monitor ongoing MPXV evolution, we performed validation against established metagenomic protocol using Illumina short-read technology at CDC^17,62^ (Methods). In total, 18 samples were sequenced by CDC and the resulting MPXV genomes were post-processed with lineages assigned using the same methods as those sequenced by our protocol (**Table 3**). 17 of 18 (94.4%) yielded the same lineage designations, with the only discrepancy due to a difference in resolution (**Table 3**; sample LC0000016: CDC genome yielded F.1 de-aliased as B.1.20.1, while our genome yielded B.1.20). Both methods yielded near-complete genomes with remarkably high median metrics for genome coverage (CDC: 99.96%, Labcorp: 99.77%) and complete annotations (CDC: 179, Labcorp: 178) of the 179 clade IIb RefSeq annotations. A more detailed examination of SNP-level resolution showed that our probe-based sequencing method had 100% concordance for both SNPs (1,136 of 1,136) and deletions (8 of 8) with the CDC metagenomic-based approach (**Table S2**). Only one of the seven insertions was a discrepant (**Table S2**), incidentally the same tandem repeat as the one detected in our biological replicates **(Table S1**, ATC repeat at positions 136,513– 136,569 in reference NC_063383.1). The near-perfect concordance and robust coverage metrics observed in these sample comparisons clearly illustrate the high fidelity afforded by both our MPXV WGS assay and the metagenomic-based assay provided by CDC.

**Table 3.**
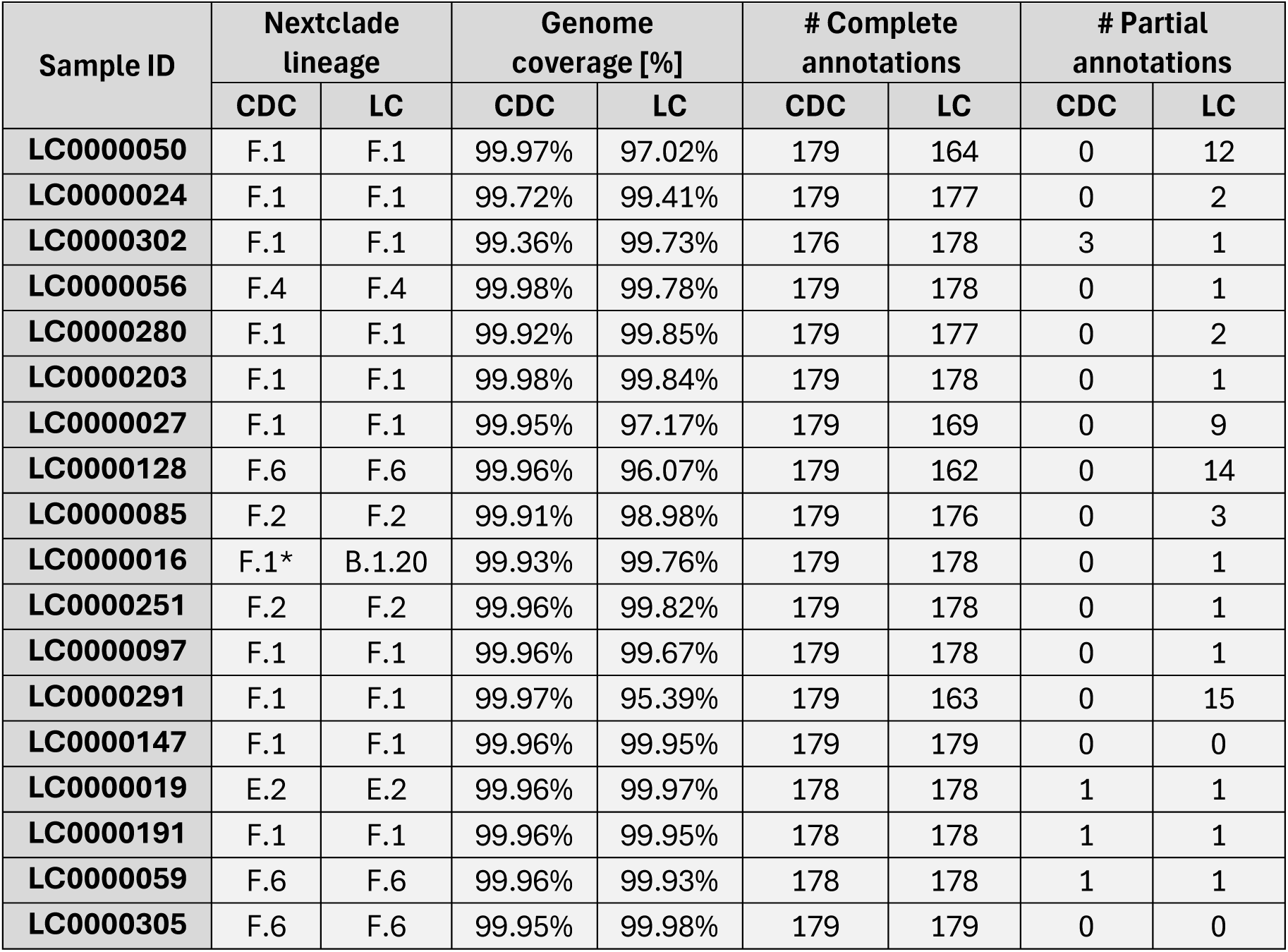
MPXV WGS accuracy assessed via parallel CDC sequencing. 18 samples were sequenced at CDC, and the resulting genomes were post-processed and compared to those sequenced at Labcorp (LC). Clade IIb lineages assigned by Nextclade^33^, genome coverage, complete gene annotations, and partial gene annotations (missing start and/or stop codon) are juxtaposed between the two sequencing methods. *F.1=B.1.20.1

| Sample ID | Nextclade lineage |  | Genome coverage [%] |  | # Complete annotations |  | # Partial annotations |  |
| --- | --- | --- | --- | --- | --- | --- | --- | --- |
|  | CDC | LC | CDC | LC | CDC | LC | CDC | LC |
| LC0000050 | F.1 | F.1 | 99.97% | 97.02% | 179 | 164 | 0 | 12 |
| LC0000024 | F.1 | F.1 | 99.72% | 99.41% | 179 | 177 | 0 | 2 |
| LC0000302 | F.1 | F.1 | 99.36% | 99.73% | 176 | 178 | 3 | 1 |
| LC0000056 | F.4 | F.4 | 99.98% | 99.78% | 179 | 178 | 0 | 1 |
| LC0000280 | F.1 | F.1 | 99.92% | 99.85% | 179 | 177 | 0 | 2 |
| LC0000203 | F.1 | F.1 | 99.98% | 99.84% | 179 | 178 | 0 | 1 |
| LC0000027 | F.1 | F.1 | 99.95% | 97.17% | 179 | 169 | 0 | 9 |
| LC0000128 | F.6 | F.6 | 99.96% | 96.07% | 179 | 162 | 0 | 14 |
| LC0000085 | F.2 | F.2 | 99.91% | 98.98% | 179 | 176 | 0 | 3 |
| LC0000016 | F.1* | B.1.20 | 99.93% | 99.76% | 179 | 178 | 0 | 1 |
| LC0000251 | F.2 | F.2 | 99.96% | 99.82% | 179 | 178 | 0 | 1 |
| LC0000097 | F.1 | F.1 | 99.96% | 99.67% | 179 | 178 | 0 | 1 |
| LC0000291 | F.1 | F.1 | 99.97% | 95.39% | 179 | 163 | 0 | 15 |
| LC0000147 | F.1 | F.1 | 99.96% | 99.95% | 179 | 179 | 0 | 0 |
| LC0000019 | E.2 | E.2 | 99.96% | 99.97% | 178 | 178 | 1 | 1 |
| LC0000191 | F.1 | F.1 | 99.96% | 99.95% | 178 | 178 | 1 | 1 |
| LC0000059 | F.6 | F.6 | 99.96% | 99.93% | 178 | 178 | 1 | 1 |
| LC0000305 | F.6 | F.6 | 99.95% | 99.98% | 179 | 179 | 0 | 0 |

### Simulation of Labcorp MPXV WGS assay demonstrates robust clade and lineage discrimination across known MPXV diversity

Our MIP-based probe panel was primarily designed to target clade IIb genomic sequences, which may result in reduced probe coverage across genomic regions that are more divergent in clades Ia, Ib, and IIa. While clades Ib and IIb have been observed in our US genomic surveillance network, other MPXV clades, including Ia and IIa, remain unobserved, likely due to their endemic circulation outside the US, lack of US cases due to travel to those areas, and the absence of zoonotic transmission domestically. This limits our ability to directly evaluate assay performance across the full spectrum of MPXV diversity. To address this gap, we employed simulation methods to assess our assay’s capacity to detect and resolve all known MPXV clades and lineages. We previously developed a simulator for our Virseq assay, which used a coverage and error model with a reference-based approach^11^. Since MPXV clades differ significantly evolutionarily and are known to harbor various structural variations^14,63^, we elected to take a different approach for simulating MPXV that modelled the probe design of our assay (Methods). The simulator developed in this study functions similarly, where the user specifies an input ‘ground truth’ MPXV genome and a depth of coverage to simulate. However, instead of simulating genomes directly, this approach generates CCS reads that are then carried through the same bioinformatics processes as in our MPXV WGS assay.

We simulated all known MPXV clades and additionally all clade IIb lineages not yet observed by our MPXV surveillance efforts with available genomes, constituting a set of four clade Ia, five clade Ib, five clade IIa, and 65 clade IIb high coverage genomes (Methods). These genomes were processed through our simulator at four different target depths of coverage in triplicate then post-processed using our MPXV bioinformatics pipeline, yielding 939 simulated genomes (**Table 4**, **Figure S8**). In general, target and simulated depths of coverage were highly similar regardless of MPXV clade, though a slight negative bias was observed in the simulated coverages (0.01 to −2.15 bias; 0.1 to - 7.2% difference from target; **Table 4**). This was anticipated, since low quality simulated reads might be removed during genome assembly, and alignment soft-clipping might also affect slight reductions in coverage depth yielded for the simulated MPXV genomes. Lastly, we observed increasing genome coverage as simulated depth of coverage increased for all clades (**Figure S9**), consistent with observations from our MPXV WGS surveillance data (**Figure 3a**). In general, we observed a slight negative shift in clade IIb genome coverage even at higher depths of coverage simulated for some input genomes (**Figures S7** and **S8**). This was likely an artifact arising from MIP selection stringency in our simulation algorithm, where probes were unable to align to regions with ambiguous nucleotides in input genomes, resulting in coverage gaps that would not occur with real clinical specimens containing complete viral sequences (Methods). Indeed, manual inspection of these genomes showed that they were lower quality (∼93.6–97.2% coverage) compared to input Clade IIb genomes in general (median=99.99%; **Supplementary Data**). Despite this limitation for some lower quality input genomes, overall, the simulator reliably generated reads closely in line with target sequencing depth and was compatible with our WGS post-processing workflow.

**Table 4.**
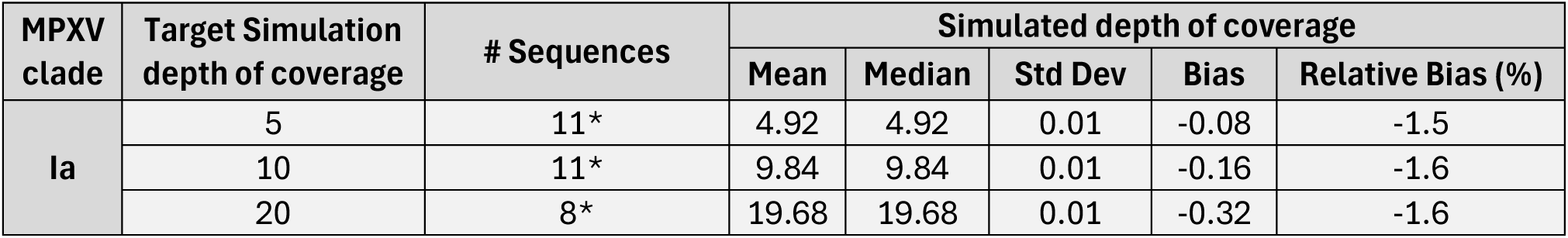
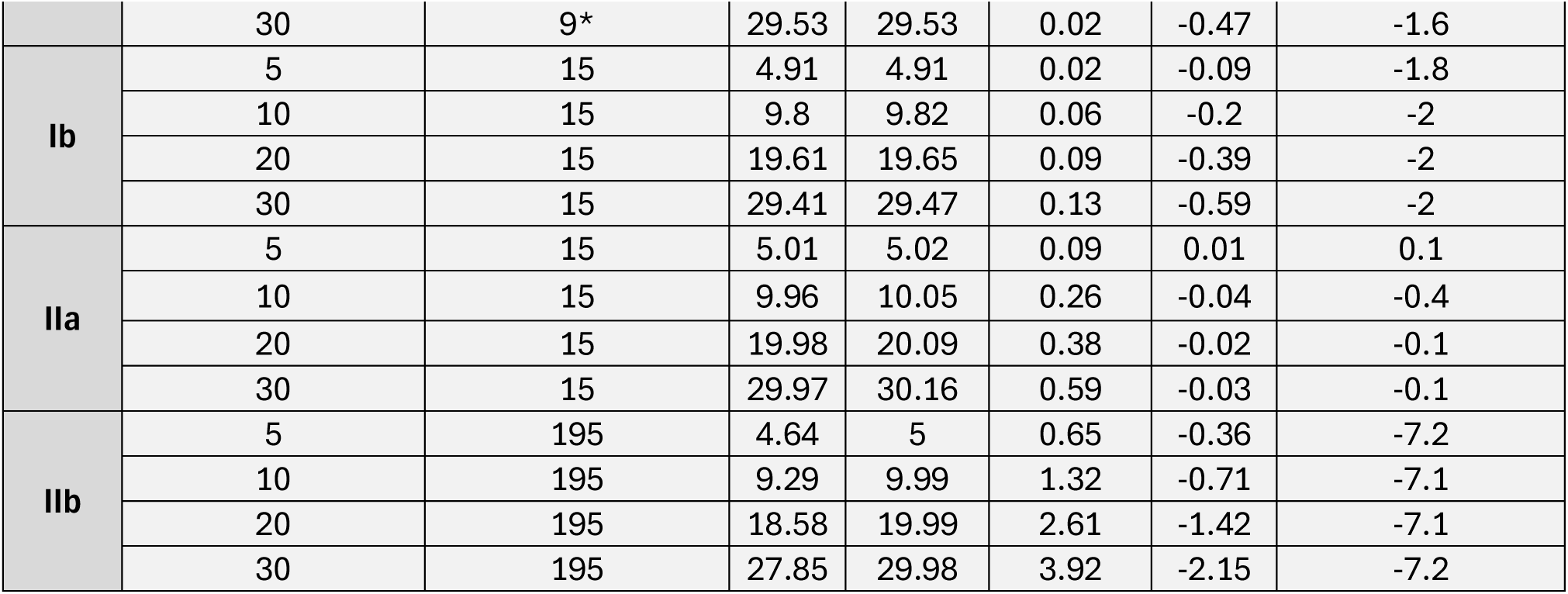
MPXV genome coverage simulation. Each section of the table describes coverage simulation results for a different MPXV clade (Ia, Ib, IIa, IIb), showing the number of sequences, target depth of coverage and the summary statistics yielded in the simulated genomes. Each clade was simulated across four different depths of coverage (5X, 10X, 20X, 30X). *Some Clade Ia simulation replicates failed bioinformatics processing (one 5X: OR943698.1; two 10X: HM172544.1; four 20X: HM172544.1, NC_003310.1, OP498046.1; three 30X: HM172544.1, NC_003310.1, OR943698.1).

| MPXV clade | Target Simulation depth of coverage | # Sequences | Simulated depth of coverage |  |  |  |  |
| --- | --- | --- | --- | --- | --- | --- | --- |
|  |  |  | Mean | Median | Std Dev | Bias | Relative Bias (%) |
| Ia | 5 | 11* | 4.92 | 4.92 | 0.01 | -0.08 | -1.5 |
|  | 10 | 11* | 9.84 | 9.84 | 0.01 | -0.16 | -1.6 |
|  | 20 | 8* | 19.68 | 19.68 | 0.01 | -0.32 | -1.6 |
|  | 30 | 9* | 29.53 | 29.53 | 0.02 | -0.47 | -1.6 |
| Ib | 5 | 15 | 4.91 | 4.91 | 0.02 | -0.09 | -1.8 |
|  | 10 | 15 | 9.8 | 9.82 | 0.06 | -0.2 | -2 |
|  | 20 | 15 | 19.61 | 19.65 | 0.09 | -0.39 | -2 |
|  | 30 | 15 | 29.41 | 29.47 | 0.13 | -0.59 | -2 |
| IIa | 5 | 15 | 5.01 | 5.02 | 0.09 | 0.01 | 0.1 |
|  | 10 | 15 | 9.96 | 10.05 | 0.26 | -0.04 | -0.4 |
|  | 20 | 15 | 19.98 | 20.09 | 0.38 | -0.02 | -0.1 |
|  | 30 | 15 | 29.97 | 30.16 | 0.59 | -0.03 | -0.1 |
| IIb | 5 | 195 | 4.64 | 5 | 0.65 | -0.36 | -7.2 |
|  | 10 | 195 | 9.29 | 9.99 | 1.32 | -0.71 | -7.1 |
|  | 20 | 195 | 18.58 | 19.99 | 2.61 | -1.42 | -7.1 |
|  | 30 | 195 | 27.85 | 29.98 | 3.92 | -2.15 | -7.2 |

Next, we assigned clades and lineages (clade IIb only) to the simulated MPXV genomes compared these to the ground truth (**Table 5**). We obtained perfect clade designation accuracy for all four MPXV clades across all simulated depths of coverage (**Table 5**). Within clade IIb, we observed perfect lineage concordance at our minimum QC threshold of 10X mean depth of coverage and above, with some diminished accuracy below our QC threshold at 5X depth (93.3%, 182 of 195) (**Table 5**). These low depth discrepancies were spread across seven different lineages and were not systematic, i.e., some of the simulated genomes for the lineage yielded the correct lineage (**Table S3**). These accuracy results support the broad, multi-clade capabilities of our MPXV WGS assay and further show the robustness of our probe-based sequencing approach.

**Table 5.** MPXV simulation accuracy. MPXV clade simulation accuracy is shown for each simulated depth of coverage, and lineage accuracy is shown when applicable (i.e., clade IIb). Genome coverage and sequencing depth distributions for all simulated genomes are provided in **Figures S7** and **S8**.

| MPXV clade | Simulated depth of coverage | # Sequences | Clade |  | Lineage |  |
| --- | --- | --- | --- | --- | --- | --- |
|  |  |  | # Correct | Accuracy [%] | # Correct | Accuracy [%] |
| Ia | 5 | 11 | 11 | 100% | N/A | N/A |
|  | 10 | 11 | 11 | 100% | N/A | N/A |
|  | 20 | 8 | 8 | 100% | N/A | N/A |
|  | 30 | 9 | 9 | 100% | N/A | N/A |
| Ib | 5 | 15 | 15 | 100% | N/A | N/A |
|  | 10 | 15 | 15 | 100% | N/A | N/A |
|  | 20 | 15 | 15 | 100% | N/A | N/A |
|  | 30 | 15 | 15 | 100% | N/A | N/A |
| IIa | 5 | 15 | 15 | 100% | N/A | N/A |
|  | 10 | 15 | 15 | 100% | N/A | N/A |
|  | 20 | 15 | 15 | 100% | N/A | N/A |
|  | 30 | 15 | 15 | 100% | N/A | N/A |
| IIb | 5 | 195 | 195 | 100% | 182 | 93.3% |
|  | 10 | 195 | 195 | 100% | 195 | 100% |
|  | 20 | 195 | 195 | 100% | 195 | 100% |
|  | 30 | 195 | 195 | 100% | 195 | 100% |

## Discussion

Our nationwide genomic surveillance effort demonstrates the successful adaptation of probe-based long-read sequencing infrastructure from SARS-CoV-2 to MPXV, establishing a robust platform for monitoring MPXV evolution and emerging variants across all US HHS regions. Leveraging our same framework that yielded 525,000 SARS-CoV-2 high-quality genomes reported to the CDC^11^, we applied this scalable system to MPXV and generated high-quality whole genome sequences from residual mpox-positive clinical specimens. This systematic approach enabled critical public health insights, including the detection of clade Ib introductions and comprehensive characterization of clade IIb lineage dynamics and APOBEC3-mediated evolution.

### Public health impact and clinical significance

The detection of clade Ib infections during our surveillance period exemplifies the critical role of genomic surveillance in identifying novel variant introductions. Since the surveillance timeframe analyzed in this study, we identified an additional clade Ib sample in California (NCBI Genbank reference PX442285.1). Swift communication of these findings to CDC within 24 hours supported rapid public health response and epidemiological investigation, which determined these to be isolated travel-associated cases rather than sustained community transmission. This detection capability is particularly significant given the global spread of clade Ib and its detection in an increasing number of countries^64,65^. Our nationwide network, spanning all 10 US HHS regions with access to residual specimens tested using the CDC Non-Variola Orthopoxvirus Real-Time PCR Test (510(k) FDA cleared) and the Labcorp Mpox (Orthopoxvirus) PCR Test^66^, including the Home Collection Kit authorized under EUA^23^, positions us to rapidly identify and track emerging MPXV variants that could pose elevated public health risks. The recent detection of local clade Ib transmission chains in the European Union^64^, along with WHO-reported community transmission in 16 countries as of November 14, 2025 (including the US)^65^, underscores the critical importance of our genomic surveillance infrastructure to rapidly detect and characterize emerging variants before they establish sustained community transmission.

The identification of large terminal deletions in a small but notable proportion of cases provides important insights into MPXV genomic plasticity during human transmission. These deletions, concentrated near or within ITRs, mirror observations from recent studies of clade IIb infections during the 2022 outbreak^46,59^. Gigante *et al*^46^ documented similar deletion patterns affecting genes within ITR regions and demonstrated that such deletions can emerge rapidly during transmission chains. Our probe-based sequencing approach proved particularly effective at detecting these structural variants, as the high coverage depth and complete genome recovery enabled clear, granular visualization of coverage dropout patterns that might be missed by lower-depth approaches. The functional implications of these deletions remain unclear. While their proximity to immune evasion genes, particularly those within the ITR regions, is intriguing, experimental studies are needed to determine whether these deletions provide selection advantages during human transmission or merely represent neutral genomic plasticity, as recently discussed by Gigante *et al*^46^.

### Assay Reproducibility and Cross-Platform Validation

The exquisite concordance observed between our probe-based sequencing method and CDC’s metagenomic approach validates the accuracy of both platforms and demonstrates that different sequencing strategies can yield equivalent results for MPXV genomic surveillance^17,62^. Both methods achieved near-complete genome coverage with comprehensive gene annotations and showed perfect concordance for SNPs and deletions. We observed slightly greater variability among biological replicates from the same patient compared to the near-perfect concordance between sequencing platforms. This suggests that minor discrepancies in biological replicates may reflect genuine intra-host viral diversity rather than technical artifacts. It is also worth noting that low-complexity tandem repeat regions, which are abundant in MPXV terminal regions, present well-documented challenges for short-read sequencing technologies due to ambiguous read mapping and alignment artifacts in repetitive sequence contexts. The long-read sequencing approach employed here offers inherent advantages in these regions, as longer reads can span repeat elements more completely, reducing ambiguity in both variant calling and structural variant detection^25,26^.

Intra-host APOBEC3 activity likely contributes to this variation, as APOBEC3 enzymes are expressed in various human tissues and can generate localized mutational signatures^59^. The substantial burden of APOBEC3-associated mutations we observed in clade IIb, with many being rare variants, supports ongoing mutagenesis during sustained human to human infections. Previous work has shown that APOBEC3-mediated editing can generate minority variants within individual infections that become fixed in subsequent transmission events^59^. Substantial intra-host MPXV diversity was also documented from data collected from the 2022 mpox outbreak in Melbourne, Australia^60^, and a study of long-term mpox infections investigated temporal and tissue resolution of intra-host MPXV diversity^61^. We hypothesize that the anatomical diversity of specimen collection sites in our dataset may capture distinct tissue microenvironments with varying APOBEC3 expression patterns, potentially contributing to the observed intra-patient genomic heterogeneity, although distinguishing this from technical variation might require deeper sequencing to characterize minority variants. This finding has important implications for understanding MPXV evolution and highlights that genomic surveillance can capture snapshots of ongoing intra-host evolutionary processes.

### Molecular Clock, APOBEC3-Driven Evolution, and Surveillance Implications

Our time-calibrated phylogenetic analysis employed a molecular clock rate consistent with Nextstrain recommendations and previous estimates for clade IIb^21,48^. However, this rate was calibrated primarily on early clade IIb outbreak data and may require revision as additional genomic data accumulates. The extensive APOBEC3-mediated mutagenesis we documented represents an acceleration of evolutionary rate compared to historical clade I and IIa viruses, which evolved primarily through zoonotic spillover with limited human-to-human transmission^21^. O’Toole *et al* demonstrated that APOBEC3 editing became the dominant mutational process in clade IIb following sustained human transmission beginning around 2016^21^, fundamentally altering MPXV’s evolutionary trajectory. As genomic surveillance data continues to accumulate, refinement of molecular clock estimates may be necessary to account for potential rate heterogeneity across different transmission contexts and geographic regions.

A critical question is whether clade Ib will follow a similar evolutionary pattern. Our detection of substantial APOBEC3 signatures in two clade Ib genomes suggests that this mutational mechanism is active during clade Ib human transmission chains, consistent with initial genomic analysis from the eastern DRC outbreak^67^. While our clade Ib sample size is limited, the observed APOBEC3 activity (80.4%) approaches that of clade IIb (90.2%), suggesting clade Ib may be undergoing similar host adaptation. Given the sustained community transmission of clade Ib documented in many countries^6^, continued spread could accelerate APOBEC3-driven evolution, potentially generating variants with altered phenotypic properties. Emerging studies from sustained clade Ib transmission settings, including genomic analyses from the eastern DRC outbreak, are beginning to characterize clade Ib APOBEC3 dynamics in greater depth^51,67^. Additionally, clade IIb lineage A transmission documented in West African countries, e.g., Sierra Leone^68^, provides a complementary context for understanding APOBEC3 evolution outside of the men who have sex with men (MSM)-associated lineage B outbreaks. Continued longitudinal sampling across diverse transmission settings will be important to determine whether clade Ib APOBEC3 signatures intensify with sustained human transmission as observed for clade IIb^21^.

The differential mutation density we observed across MPXV genomic regions provides insights into evolutionary constraints. Our analysis revealed significantly lower SNP density in the central conserved region compared to variable regions (V1 and V2), indicating purifying selection acting to maintain essential viral functions encoded in the core genome. This is consistent with decades of poxvirus research demonstrating that the central genomic region encodes essential replication machinery and structural proteins under strong evolutionary constraint, while terminal variable regions encode immunomodulatory and host-range factors subject to more rapid diversification^12,14,69^. This conserved genomic architecture is a defining feature of poxviruses broadly and has been exploited for diagnostic and therapeutic target selection^12^. Notably, the majority APOBEC3-associated SNPs showed even stronger regional differentiation, with significantly reduced APOBEC3 activity in the conserved core. This suggests that while APOBEC3 generates mutations genome-wide, deleterious mutations in functionally constrained regions are rapidly purged during transmission. This finding contrasts with prior reports describing relatively uniform APOBEC3 mutation distributions across the MPXV genome^21^, and may reflect the larger, more geographically diverse, and more temporally extended dataset analyzed here, which may have greater power to detect differential selective pressures across genomic regions. Alternatively, differences in analytical approaches, reference genome selection, or the specific genomic partitioning used may contribute to this discrepancy, and further investigation with standardized methods across datasets will be important to reconcile these observations. This has practical implications for diagnostic assays, vaccine design, and antiviral drug targets, as genomic regions under strong selective constraint are less likely to accumulate escape mutations. Our recent *in silico* investigation of MPXV diagnostic assays showed that PCR designs targeting genes in the conserved core encountered few mutations, such as E9L (DNA polymerase) and F3L (immune evasion protein), both of which reside in the conserved core. While the hemagglutinin-like protein (HA) also showed few mutations in primer target regions in our analysis, it is worth noting that HA is known to be more variable among poxviruses broadly^70^, and caution is warranted when considering it as a universal diagnostic target across divergent clades^70^. However, some genes in the terminal regions are also valuable diagnostic targets due to clade-specific deletions, such as C3L (complement-bind protein, clade I specific) and G2R (envelope protein, clade II specific)^71^, both of which happened to show few mutations in primer target regions in our *in silico* analysis as well^70^.

The elevated mutation rate in V1 compared to V2, even after controlling for APOBEC3 activity, is an intriguing observation. However, since our analysis was limited to clade IIb lineage B genomes sampled over approximately one year, we are cautious about drawing broad mechanistic conclusions. Whether this asymmetry reflects features specific to this lineage and time period, or represents a more general biological property of MPXV replication, would require analysis across additional clades, lineages, and poxviruses. Importantly, after masking the ITRs, APOBEC3-associated rates in V1 and the conserved core no longer differed significantly, while V2 retained the highest density, indicating that the apparent V1 enrichment in the unmasked analysis was largely driven by APOBEC3 mutations concentrated in the V1 ITR.

These findings have practical implications for surveillance, cadence, and scope. The rapid accumulation of APOBEC3 mutations and emergence of distinct lineages necessitate continuous monitoring to track evolving transmission dynamics. Our simulation analysis demonstrated that our probe-based assay maintains robust performance across all known MPXV clades and clade IIb lineages, even for variants not observed in the US. This broad specificity indicates that our MIP design can tolerate substantial sequence divergence, providing confidence that our WGS assay will successfully capture future MPXV variants arising through APOBEC3-driven evolution or recombination events. The flexibility of probe-based enrichment, which functions through hybridization rather than exact sequence matching, offers advantages over amplicon-based approaches that may fail when mutations accumulate in primer binding sites.

### Broader Applications and Future Directions

Addressing the threat of emerging and re-emerging pathogens requires coordinated efforts between public health agencies, diagnostic companies, and research institutions to build resilient surveillance infrastructure. The present study is an example of this kind of partnership. CDC and Labcorp combined CDC expertise in poxvirus genomics with the nationwide diagnostic footprint of Labcorp. This synergistic effort resulted in rapid, high-coverage MPXV surveillance in the U.S. Our successful deployment of probe-enrichment-based genomic surveillance for both SARS-CoV-2^11^ and MPXV in this study demonstrates the versatility and scalability of this platform for emerging infectious disease monitoring. The modular nature of MIP design enables rapid assay development for novel pathogens, typically requiring only reference genome sequences and 2–3 weeks for probe synthesis. We have tested this approach to characterize additional pathogens of clinical interest, including complete genome sequences from multiple human adenovirus serotypes (unpublished data). This rapid response capability positions our infrastructure to address future public health threats, including the ongoing concerns regarding highly pathogenic avian influenza A(H5N1) spillover into mammalian populations and potential human adaptation^72^. The recent detection of H5N1 in dairy cattle and sporadic human infections highlights an urgent need for expanded genomic surveillance capacity^73,74^. Our existing laboratory network, sample collection infrastructure, and bioinformatics pipelines could be rapidly adapted to H5N1 surveillance by designing influenza-specific MIP panels targeting all eight genomic segments (∼13.5 kb total). The modular nature of MIP designs would enable simultaneous coverage of multiple H5N1 clades while maintaining capacity to detect reassortment events. Given the segmented nature of influenza genomes^75^, our probe-based enrichment approach could offer advantages over amplicon methods that require careful primer design to avoid bias across segments^76^. Our existing respiratory disease surveillance networks coupled with residual specimens from our influenza PCR testing would provide sample streams analogous to our MPXV genomic surveillance model.

Beyond respiratory viruses and poxviruses, this platform could address surveillance gaps for other priority pathogens including arboviruses (dengue, chikungunya, Zika), novel coronaviruses, and bacterial pathogens of concern. The integration with routine diagnostic testing, which generates residual specimens at scale without additional patient burden, provides a sustainable model for ongoing surveillance. Our probe-based approach offers advantages for pathogens present at low viral loads or in complex sample matrices, as the enrichment step enhances target recovery compared to metagenomic sequencing alone. Future enhancements could include multiplexed MIP panels targeting multiple pathogens simultaneously (e.g., influenza, RSV, SARS-CoV-2, and MPXV in a single enrichment reaction), facilitating syndromic surveillance adaptable to seasonal and geographic variation in pathogen circulation.

An important application of genomic surveillance data not fully explored in this study is transmission network analysis and contact tracing. High-resolution genomic data enables reconstruction of transmission chains and identification of epidemiological links that may not be apparent through traditional contact investigations alone. The relatively low mutation rate of MPXV compared to RNA viruses presents both challenges and opportunities for genomic epidemiology: while fewer mutations accumulate between linked cases (potentially limiting resolution), the APOBEC3-mediated mutations we documented provide molecular markers that can distinguish transmission clusters and identify introductions from different source populations. Integration of our genomic surveillance data with metadata on exposure history, geographic location, and social network information could enable more sophisticated transmission network analyses. Such analyses have proven valuable for other pathogens, including SARS-CoV-2^77,78^ and tuberculosis^79,80^, where genomic data combined with epidemiological investigation revealed unexpected transmission pathways and informed targeted interventions. For MPXV, genomic-informed contact tracing would be particularly valuable for distinguishing sustained community transmission from multiple independent introductions, as we demonstrated for clade Ib cases. Future directions should include development of analytical frameworks that integrate genomic distances, temporal data, and epidemiological metadata for real-time transmission network reconstruction, enabling public health authorities to implement more precise control measures.

### Study limitations

A few limitations should be considered when interpreting these findings. First, our surveillance captures only patients seeking medical care and tested for mpox, which may underrepresent asymptomatic or mild infections and introduce demographic biases. The demographic distribution of detected cases likely reflects both true transmission patterns and differential healthcare-seeking behavior or testing practices. Second, geographic coverage, while spanning all US HHS regions, was concentrated in high-population areas and major ports of entry, potentially missing transmission dynamics in rural or underserved communities. Third, the small number of clade Ib infections (n=3) limited our ability to perform phylogenetic APOBEC3 analysis or draw definitive conclusions about clade Ib evolutionary dynamics in the US context. While these samples provided important proof-of-concept for our detection capabilities, larger case series from sustained transmission settings would be needed to fully characterize clade Ib APOBEC3 signatures and evolutionary patterns. More broadly, our clade IIb findings are based on US surveillance data, which predominantly reflects lineage B transmission in MSM networks. These results may not fully generalize to other epidemiological contexts, including endemic settings in Africa or clade IIb lineage A transmission documented in West African countries, where distinct demographic, behavioral, and immunological factors may influence evolutionary dynamics differently.

Technical limitations include the challenges of accurately sequencing tandem repeat regions, as evidenced by insertion call discordance in specific repeat regions across both biological replicates and cross-platform comparisons. These low-complexity regions remain challenging for all current sequencing technologies and may harbor genuine biological variation that is difficult to resolve. Our deletion detection approach, while effective, relied on manual inspection of coverage profiles and may have missed smaller structural variants below our detection threshold. The simulation analysis, while comprehensive, was limited by the requirement for high-quality input genomes and may not fully capture assay performance for highly diverged or structurally complex variants that have yet to emerge.

Finally, our study focused on genomic surveillance and did not include phenotypic characterization of detected variants. The functional implications of observed mutations, particularly in genes within deleted regions or those harboring multiple APOBEC3-associated changes, remain to be determined through experimental validation. Integration of genomic surveillance data with clinical outcomes, transmission chain investigations, and viral phenotyping would provide a more complete picture of MPXV evolution and its public health implications.

## Conclusions

This study establishes a robust, scalable genomic surveillance platform for MPXV by leveraging probe-based long-read sequencing infrastructure originally developed for SARS-CoV-2^11^. Across 326 high-quality whole genome sequences spanning all US HHS regions, we successfully detected clade Ib introductions and comprehensively characterized clade IIb evolution, including dynamic lineage shifts, extensive APOBEC3-mediated mutagenesis with regional genomic variation, and rare large terminal deletions. Our platform demonstrated exceptional accuracy through concordance with CDC metagenomic sequencing, reproducibility across biological and technical replicates, and robust performance across all known MPXV clades and lineages via simulation analysis. These findings underscore the critical role of genomic surveillance in detecting emerging variants, understanding viral evolution during sustained human transmission, and informing public health responses. Realizing the full potential of such surveillance infrastructure requires sustained collaboration among public health authorities, academic research institutions, and diagnostic industry partners to integrate regulatory frameworks, laboratory capacity, bioinformatics expertise, and epidemiological investigation into coordinated response networks. The modular probe-based design and demonstrated success across multiple pathogens (SARS-CoV-2 and MPXV) position this infrastructure to rapidly adapt to future emerging infectious disease threats, including H5N1 and novel zoonotic spillovers, exemplifying how investment in genomic surveillance capacity yields dividends across diverse public health challenges.

## Data Availability

Figures S1-S8 and Tables S1-S3 are available in the Supplementary Materials. Full simulation results, including source genome details and simulated genome parameters and outcomes are available in the Supplementary Data. De-identified Labcorp MPXV genome analysis results, metadata, limited demographic data, and replicate information are available in the Supplementary Data. The genome sequences generated in this study were deposited in the Pathoplexus database (pathoplexus.org; https://doi.org/10.62599/PP_SS_2806.1).

## List of abbreviations

APOBEC3: Apolipoprotein B mRNA Editing Enzyme, Catalytic Polypeptide-like 3
BAM: Binary Alignment Map
bp: base pairs
C: Central conserved region
CCS: Circular consensus sequence
CDC: Centers for Disease Control and Prevention
Ct: Cycle threshold
DNA: Deoxyribonucleic acid
DRC: Democratic Republic of the Congo
EUA: Emergency Use Authorization
FDA: Food and Drug Administration
GISAID: Global Initiative on Sharing All Influenza Data
GFF: General Feature Format
HHS: Health and Human Services
IQR: Interquartile range
ITR: Inverted terminal repeat
kb: kilobases
MIP: Molecular Inversion Probe
MPXV: Mpox virus
MSM: Men who have sex with men
NCBI: National Center for Biotechnology Information
NGS: Next-generation sequencing
PCR: Polymerase chain reaction
PHEIC: Public Health Emergency of International Concern
QC: Quality control
R_0_: Basic reproduction number
RNA: Ribonucleic acid
SARS-CoV-2: Severe acute respiratory syndrome coronavirus 2
SMRT: Single Molecule, Real-Time
SNP: Single nucleotide polymorphism
US: United States
V1: Left variable region
V2: Right variable region
VCF: Variant Call Format
WGS: Whole genome sequencing
WHO: World Health Organization

## Declarations

### Ethics approval and consent to participate

This study was determined to be exempt from IRB review as it involved retrospective analysis of de-identified residual clinical specimens collected during routine diagnostic testing. No additional patient contact or informed consent was required.

### Availability of data and materials

**Figures S1-S8** and **Tables S1-S3** are available in the **Supplementary Materials**. Full simulation results, including source genome details and simulated genome parameters and outcomes are available in the **Supplementary Data**. De-identified Labcorp MPXV genome analysis results, metadata, limited demographic data, and replicate information are available in the **Supplementary Data**. The genome sequences generated in this study were deposited in the Pathoplexus database (pathoplexus.org; https://doi.org/10.62599/PP_SS_2806.1).

### Authors’ contributions

Labcorp mpox WGS assay development and conceptualization – H.N.B., J.M., B.M.N., J.D.W., A.B.H., D.B., S.E.D., L.K.I. Mpox WGS sample collection, metadata collation, and sequencing – H.N.B., Q.Zh., J.M., N.J.H., N.B., S.E.D., L.K.I. CDC mpox WGS and analysis – J.Chai., C.H., D.M., and C.M.G. Manuscript supervision and administration – H.N.B. and L.K.I. Manuscript conceptualization and methodology – H.N.B., Q.S., K.S., Q.Zh., and L.K.I. Data processing and curation – H.N.B. and Q.Zh. CLC configuration – Q.Ze., K.K., and J.Chap. Formal analysis and generation of figures and tables – H.N.B., Q.S., K.S., and Q.Zh. Formal analysis review – H.N.B., Q.S., K.S., Q.Zh., and L.K.I. Original manuscript draft preparation – H.N.B. Manuscript review and editing – all authors. Approval of final manuscript – all authors.

## Acknowledgements

We would like to acknowledge the dedicated members of our IT infrastructure team who continue to be instrumental in maintaining the resources needed for this surveillance effort. We also thank the technicians and technologists who processed mpox PCR testing at Labcorp. We would also like to extend our gratitude to the Molecular Loop Biosciences® team who were instrumental in the development of the Labcorp mpox WGS assay. Lastly, we would like to thank senior leadership at Labcorp their continued support for public health surveillance efforts in the post-COVID-19 pandemic era.

