## Supplementary Materials for "Nationwide Mpox Genomic Surveillance Reveals Clade Ib Introductions, APOBEC3-Driven Evolution, and Terminal Deletions"


| Page # | Supplementary Figure/Table |
|---|---|
| 2 | **Figure S1.** VAC1 Ct value distribution of sequenced mpox-positive specimens. |
| 3 | **Figure S2.** Analysis of genome coverage outliers. |
| 4 | **Figure S3.** Visualization of genomic deletions detected across ten samples with coverage anomalies analyzed in Figure S2. |
| 5 | **Figure S4.** Iterative refinement of large-deletion breakpoints in the 10 samples with dropout |
| 7 | **Figure S5.** MPXV phylogenetic tree showing APOBEC3-like mutation patterns. |
| 8 | **Figure S6.** Mean depth of coverage analysis across MPXV conserved (C) and variable regions (V1, V2). |
| 9 | **Figure S7.** MPXV regional SNP densities with ITRs masked. |
| 10 | **Figure S8.** Simulated mean depth of coverage across MPXV clades. |
| 11 | **Figure S9.** Relationship of simulated coverage metrics across MPXV clades. |
| 12 | **Table S1.** Mutation-level concordance of biological and technical replicates. |
| 13 | **Table S2.** Mutation-level concordance of Labcorp and CDC sequence MPXV genomes. |
| 14 | **Table S3.** Clade IIb simulation lineage accuracy results. |

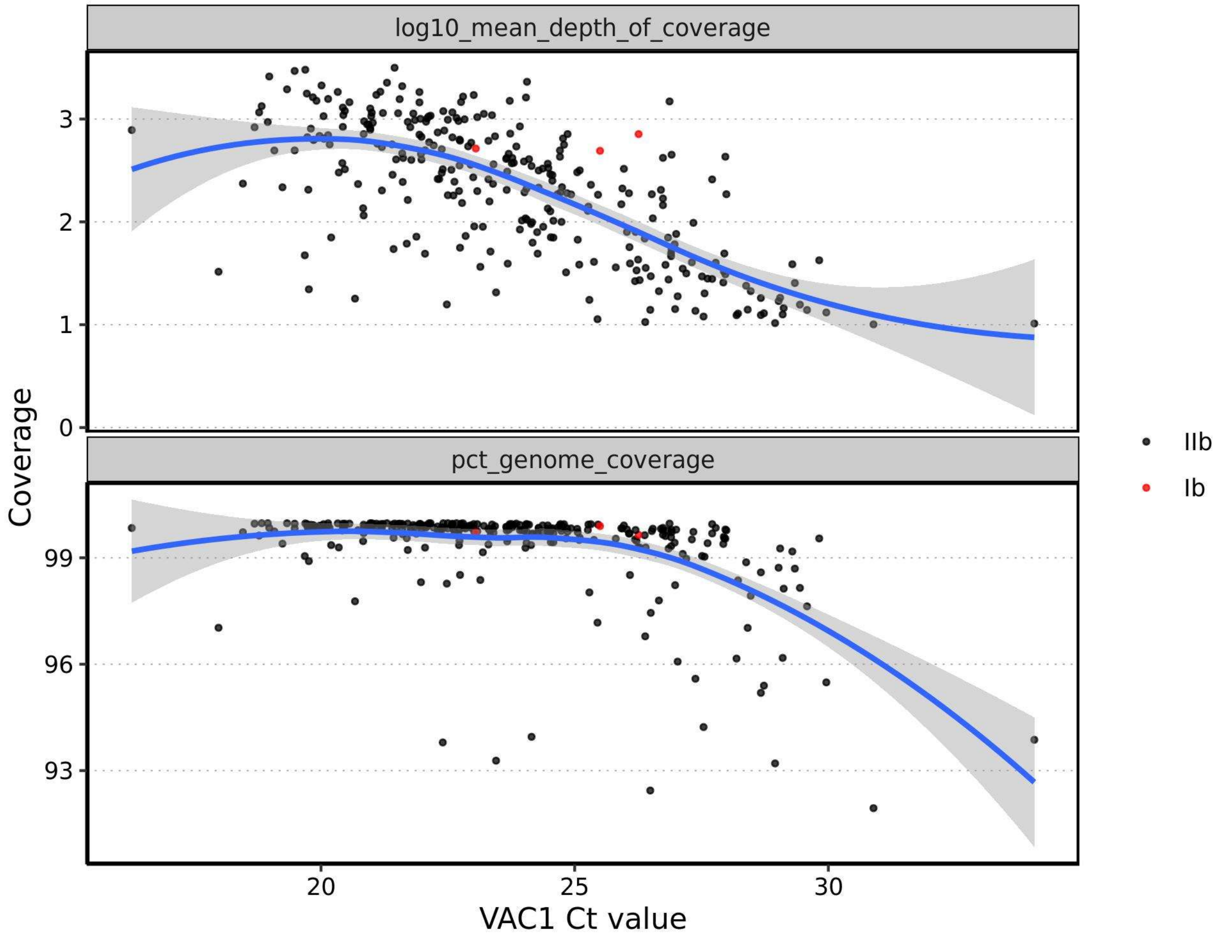


**Figure S1. VAC1 Ct value distribution of sequenced mpox-positive specimens.** VAC1 Ct values are plotted versus the mean depth of coverage (log10; top) and the genome coverage (%, bottom) with each data point representing a sequenced genome from an mpox-positive specimen. Data points are colored by mpox virus (MPXV) clade (Ib: red, IIb: black) and a loess curve for each coverage metric is shown in blue with grey fill indicating the 95% confidence interval.

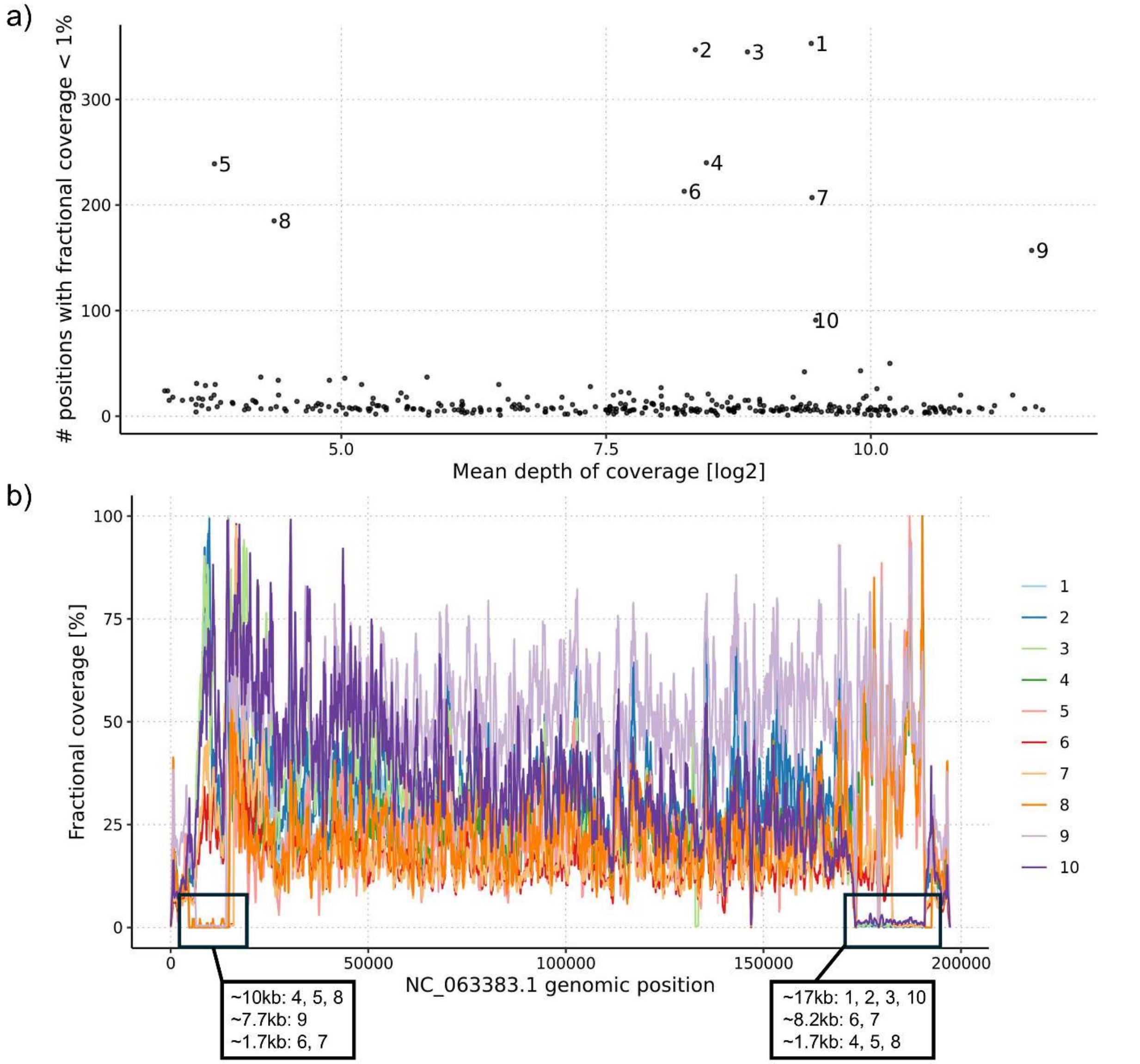


**Figure S2. Analysis of genome coverage outliers. (a)** Sequenced genomes are plotted by their mean depth of coverage (log2, x-axis) and number (#) of genomic positions with fractional coverage < 1% (y-axis). Fractional coverage is defined as the coverage at a genomic position divided by the maximum coverage detected across all genomic positions for that sample. Ten outliers with >75 positions with <1% fractional coverage are denoted in order of most to fewest positions. **(b)** Fractional coverage [%] profiles of the ten outlier samples from 5' (left) to 3' (right), with long stretches of <1% fractional coverage (i.e., deletions) marked by black boxes that include the lengths and corresponding samples annotated.

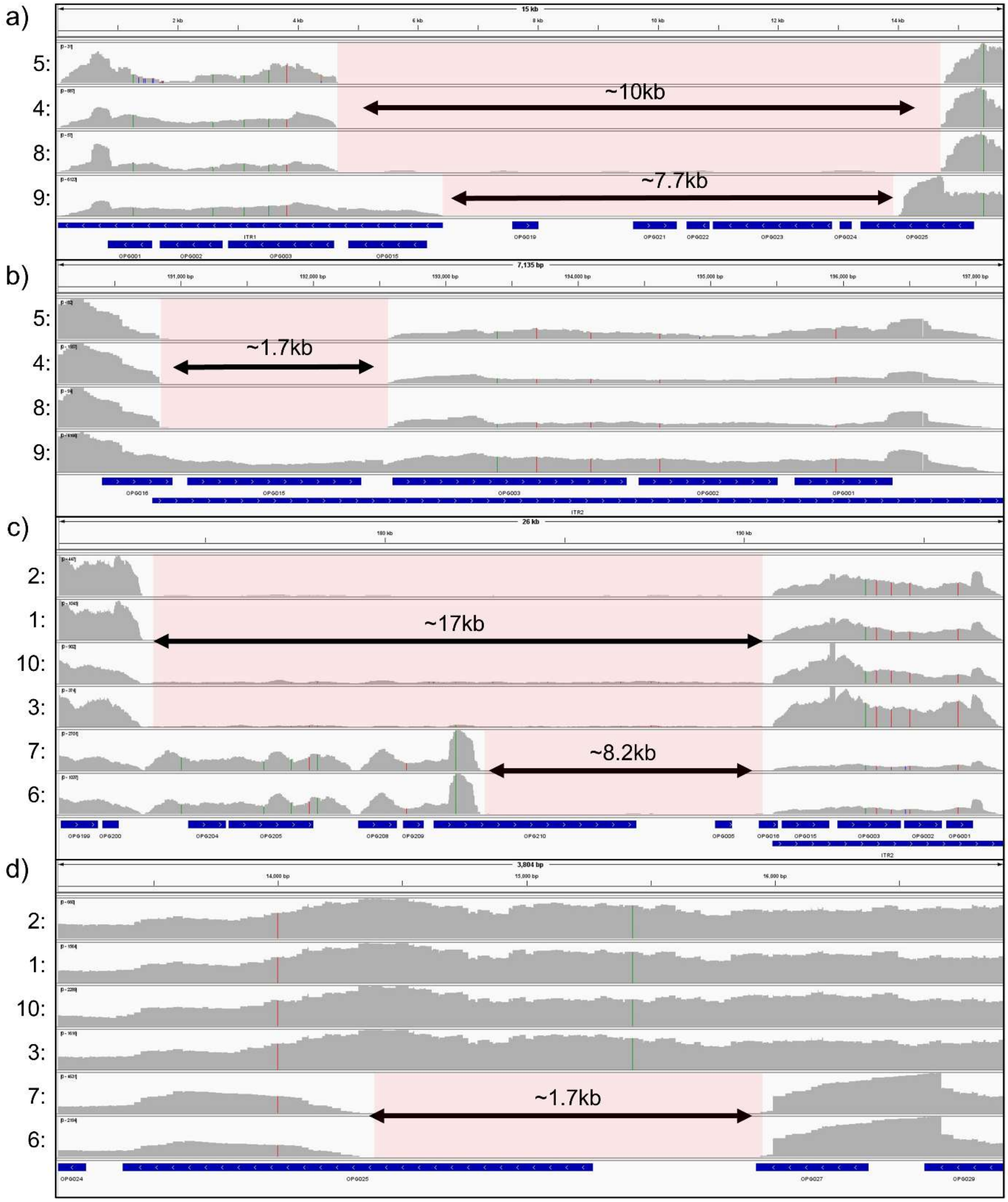


**Figure S3. Visualization of genomic deletions detected across ten samples with coverage anomalies analyzed in Figure S2.** Each row shows the coverage track of a sample, and deletions are indicated by red boxes with double-sided arrows and approximate lengths in kilobases (kb). Panels (a) and (d) show deletions in 5' proximal genomic regions, while (b-c) show deletions in 3' proximal genomic regions. Samples are grouped by common deletions detected in (a-b) and (c-d).

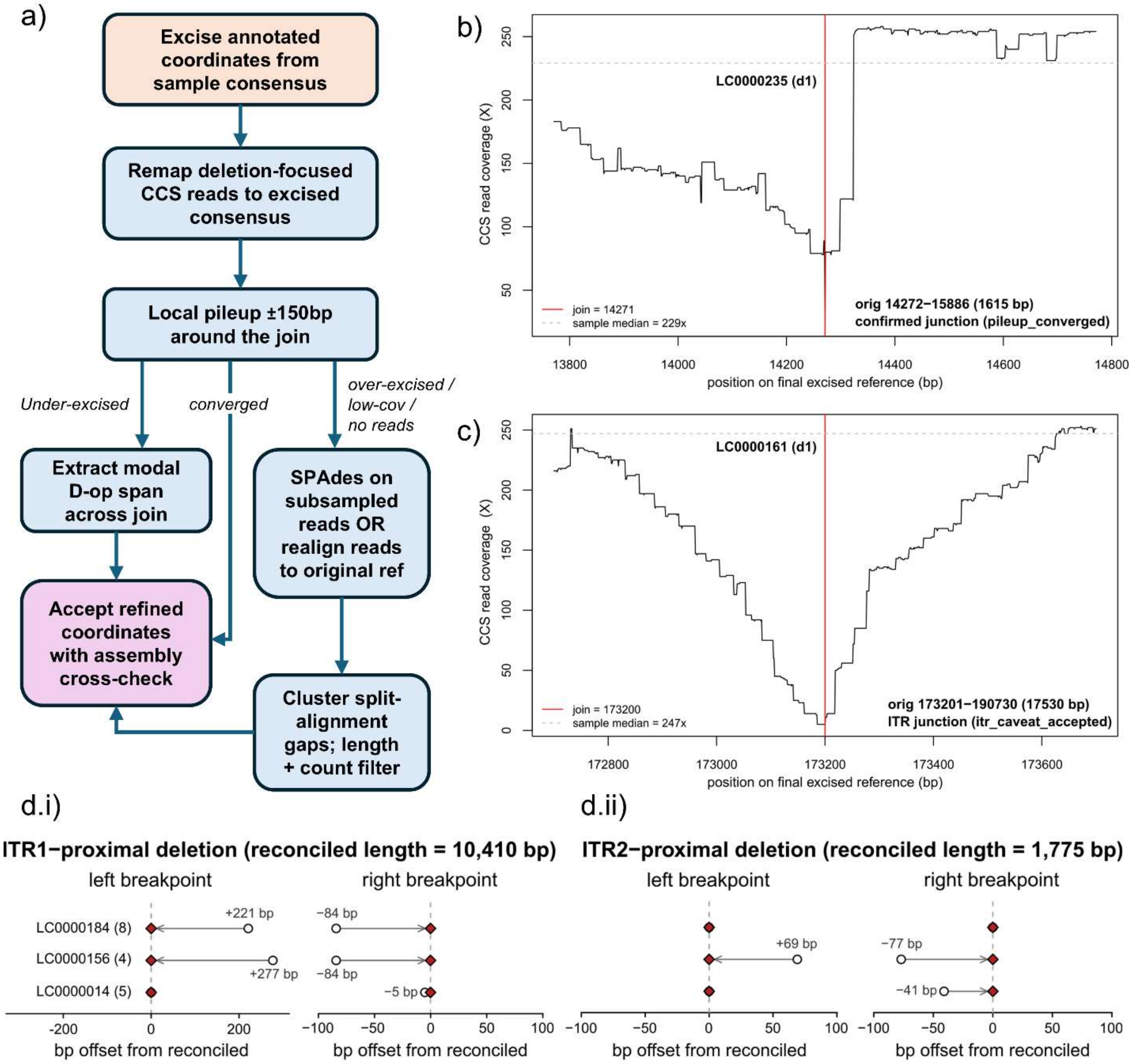


**Figure S4. Iterative refinement of large-deletion breakpoints in the 10 samples with dropout.** Initial deletion coordinates from IGV inspection (**Figure S3**) were refined at base-pair resolution against per-sample excised references using an iterative alignment–pileup routine, cross-checked against a full-sample de novo assembly. **(a)** Refinement workflow where each putative deletion was excised from the sample consensus. Deletion-focused CCS reads (±5 kb of either breakpoint, plus all unmapped reads) were remapped to the excised reference with minimap2 (-x map-pb), and a local pileup spanning ±250 bp of the join classified the junction as *converged*, *under-excised*, or *over-excised*. Under-excised joins were refined from the modal D-op span across the join; over-excised or low-coverage joins were routed to a de novo assembly path (subsampled reads or SPAdes contigs remapped to the original reference, breakpoints from split-alignment reference gaps clustered at ±200 bp). A full-sample SPAdes --isolate assembly provided an independent cross-check. **(b)** CCS coverage across ±500 bp of the join for a representative confirmed junction (LC0000235 d1; 1,615 bp, ITR1; evidence_class = pileup_converged). The symmetric coverage attenuation flanking the join is the expected signature of a correctly excised deletion, where probes straddling the deletion boundary have one arm annealing to deleted sequence and fail to circularize, producing a dropout band ~1 insert length wide (~675bp) on each side. **c)** Same view for an ITR-proximal deletion (LC0000161 d1; 17,530 bp; evidence_class = itr_caveat_accepted). The ITR-caveat class reflects that near-identical inverted terminal repeats prevent independent assembly recovery, so the coordinate calls rest on the excised-reference read alignment alone, yielding similar coverage geometry at the junction. **(d.i)** and **(d.ii)** sub-panels describe same-donor breakpoint reconciliation for the biological triplicates LC0000014 (5), LC0000156 (4), and LC0000184 (8). Sub-panels show the ITR1- and ITR2-proximal deletions; within each, left and right breakpoints are plotted side-by-side. Each sample's raw IGV coordinate was projected onto the last position shared across the three donor consensuses within a 1.5 kb flank (anchor: LC0000014). Open circles indicated the raw coordinates with signed offsets, filled red diamonds are the reconciled coordinates (x = 0, identical across samples).

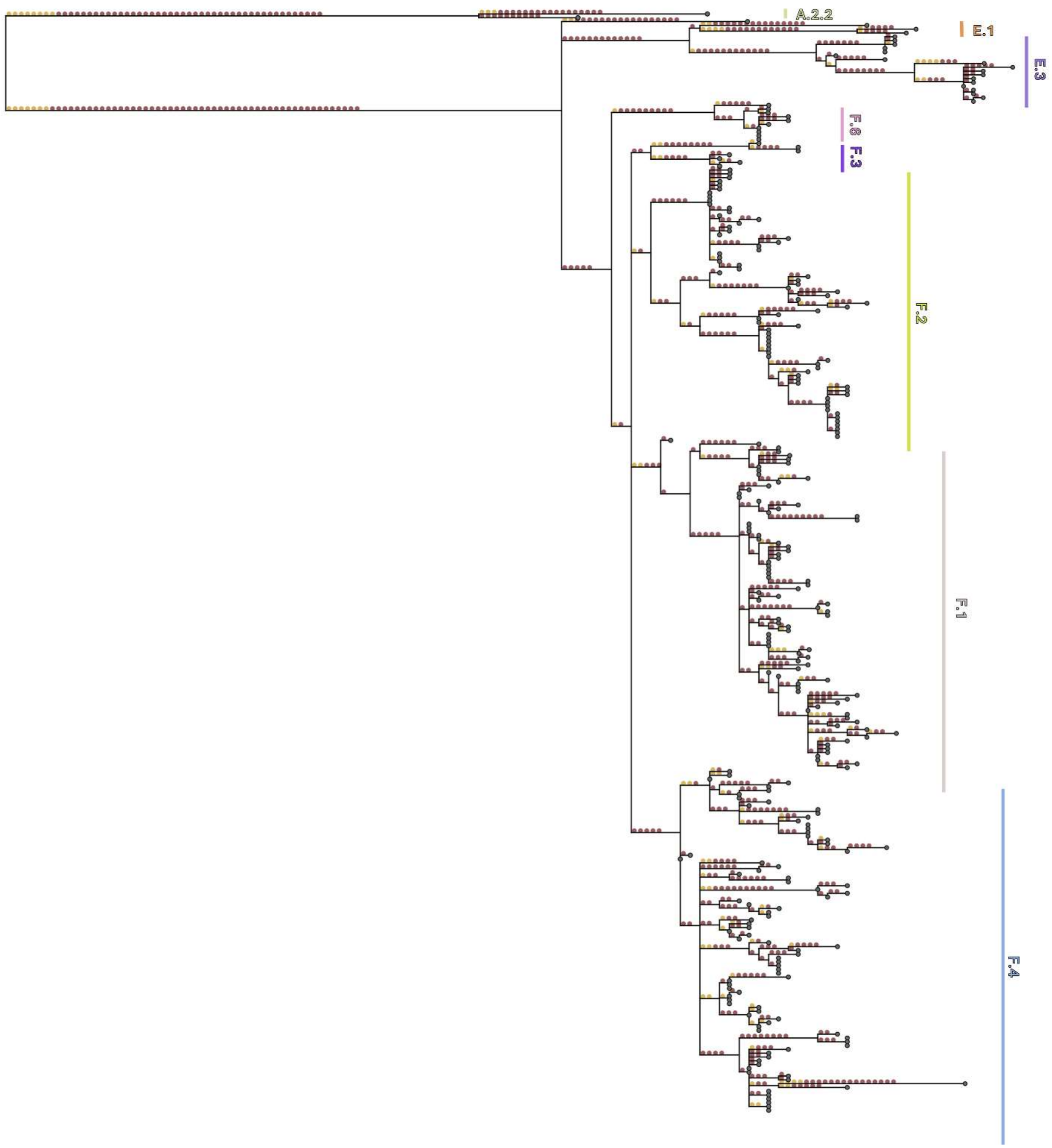


**Figure S5. MPXV phylogenetic tree showing APOBEC3-like mutation patterns.** Branch mutations are shown as circles: red for APOBEC3-like mutations (TC→TT, GA→AA) and yellow for other mutations. Major lineages are grouped and highlighted with matching color keys. Only clusters with more than 2 samples are annotated with text, including A.2.2, E.1, E.3, F.6, F.3, F.2, F.1, and F.4 from top to bottom of the phylogenetic tree.

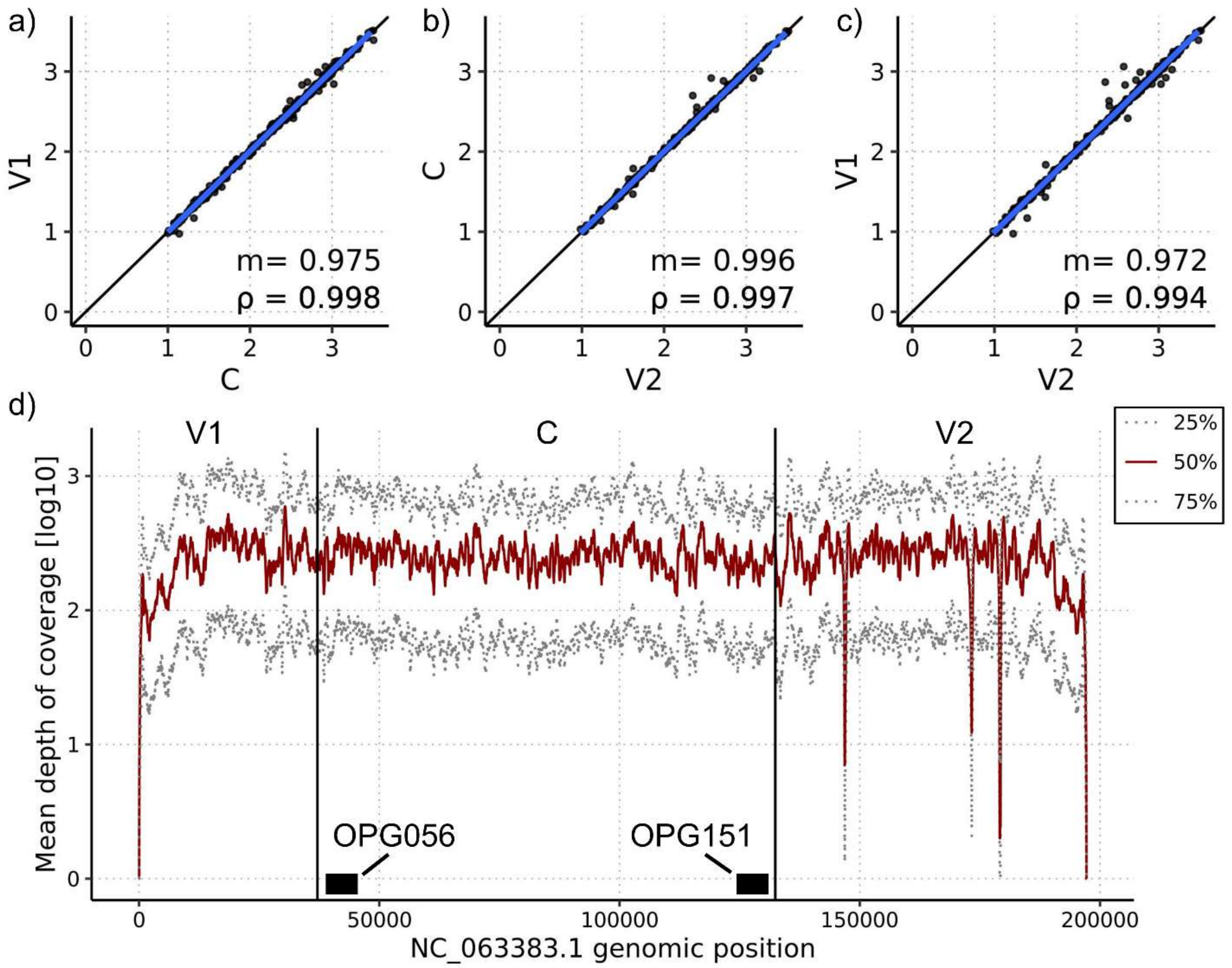


**Figure S6. Mean depth of coverage analysis across MPXV conserved (C) and variable regions (V1, V2). (a-c)** Pairwise comparisons of mean depths of coverage (log10) for each sequenced MPXV genome between regions (V1, C, V2). A linear fit is shown as a blue line and y=x as a black line, with slope (m) and spearman correlation (ρ) shown on the bottom right of each panel. **(d)** Per-base mean depth of coverage (log10) for quartiles across the MPXV genome (grey dotted: 25%, solid red: 50%, gray dotted: 75%). Regions (V1, C, V2) are labelled and demarcated by vertical black lines. The C region encompasses all RefSeq annotated genes from OPG056 to OPG151, which are shown at the 5' and 3' proximal ends of the C region, respectively.

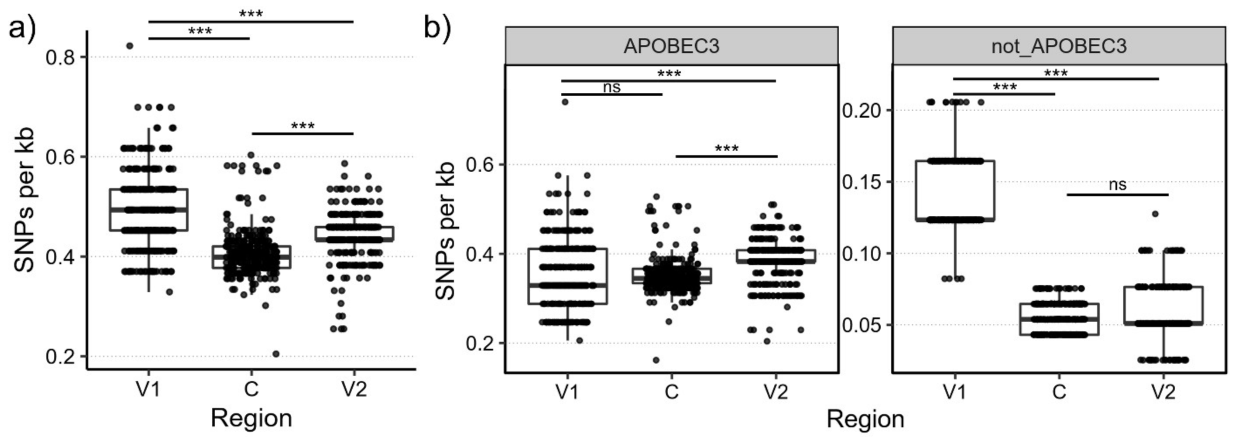


**Figure S7. MPXV regional SNP densities with ITRs masked.** SNPs per kb across conserved (C) and variable regions (V1, V2) overall (a) and stratified by APOBEC3 association (b). The C region encompasses all RefSeq annotated genes from OPG056 to OPG151, V1 encompasses gene 5' of OPG056, and V2 encompasses genes 3' of OPG151. ITRs in V1 (ITR1) and V2 (ITR2) were masked and thus removed from this analysis. ****P*<0.001; ns=not significant.

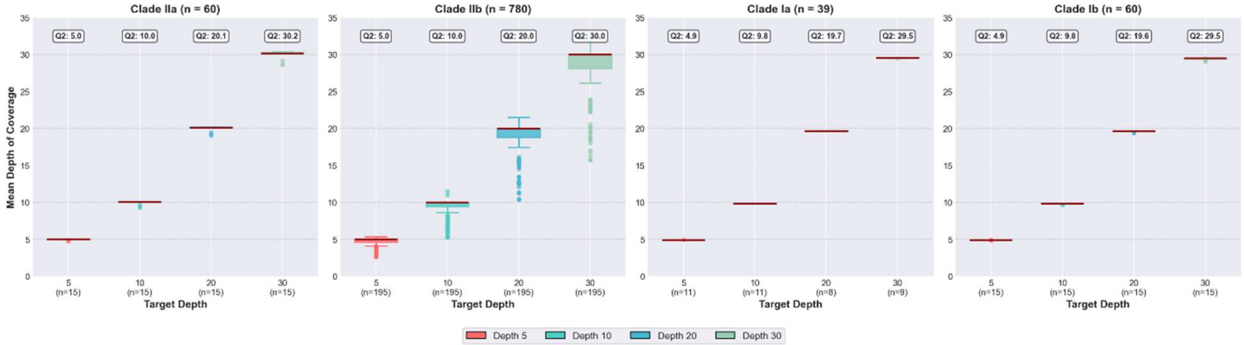


**Figure S8. Simulated mean depth of coverage across MPXV clades.** Box plots display the distribution of simulated mean depth of coverage for sequences at four target depths (left-to-right in each panel: red: 5X, cyan: 10X, blue: 20X, green: 30X) across four major MPXV clades (left-to-right panels: Clade IIa, IIb, Ia, Ib). The total number of sequences simulated for each clade is shown in each panel title. Above each box plot is the median (Q2) simulated mean depth of coverage.

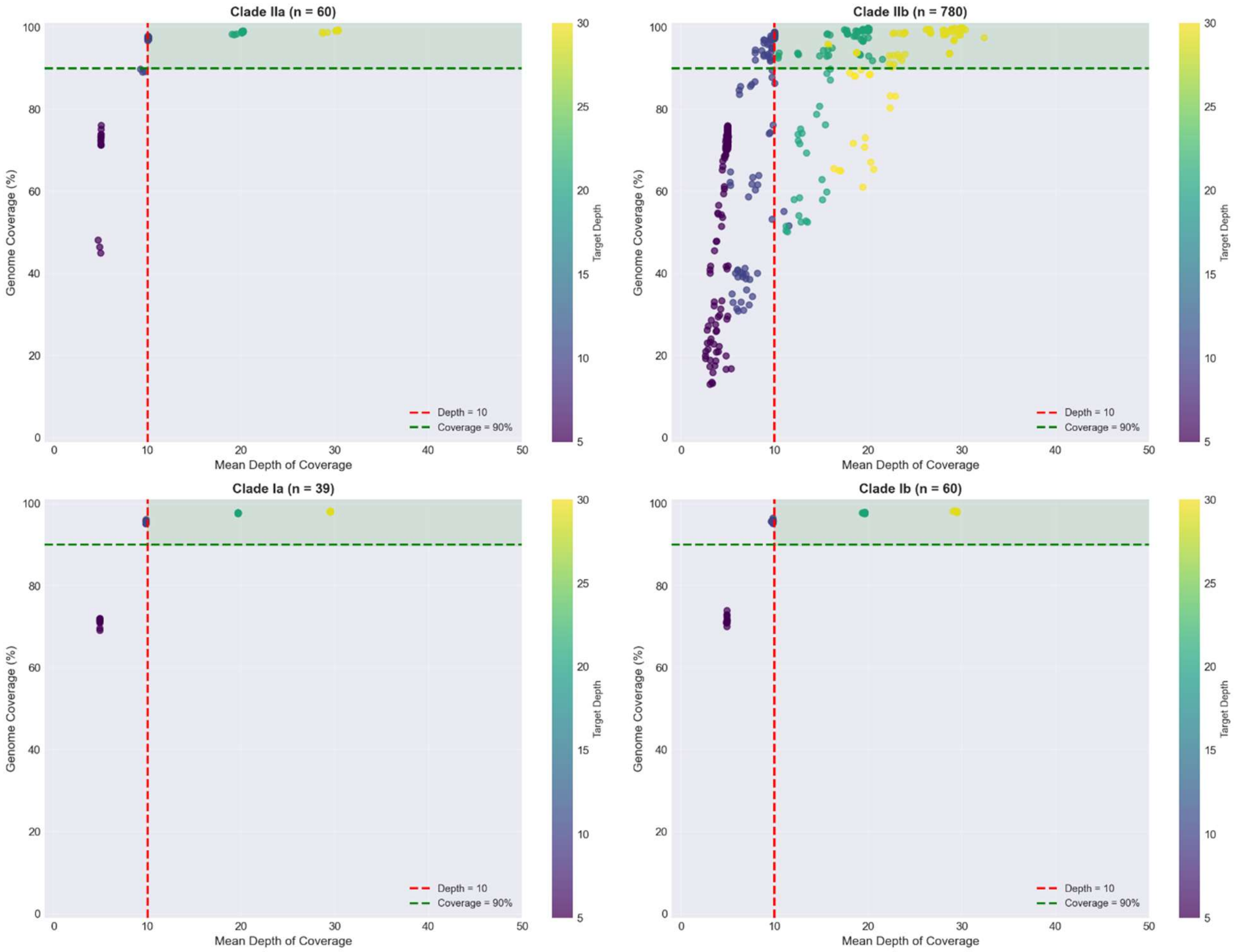


**Figure S9. Relationship of simulated coverage metrics across MPXV clades.** Scatter plots show simulated mean depth of coverage (x-axis) versus % genome coverage (y-axis) for all simulated sequences. Each data point represents a simulation sequence, colored by target sequencing depth (purple: 5X, blue: 10X, cyan: 20X, yellow: 30X). Dashed lines indicate minimum quality control thresholds imposed for sequence analysis in this study (vertical red: minimum depth of coverage, horizontal green: minimum % genome coverage). Each panel shows simulation results for a clade with the number of simulated sequences shown (top-left: IIa, top-right: IIb, bottom-left: Ia, bottom-right: Ib).

| Replicate | | | | Substitutions | | | Deletions | | | Insertions | | |
|---|---|---|---|---|---|---|---|---|---|---|---|---|
| Type | ID | # Samples | # Genes | Total | Concordant # | Concordant % | Total | Concordant # | Concordant % | Total | Concordant # | Concordant % |
| Biological (same patient, different specimen) | **bio1** | **3** | **133** | 28 | 28 | 100% | 0 | 0 | N/A | 1 | 0 | 0% |
| | **bio2** | **2** | **174** | 74 | 73 | 98.6% | 0 | 0 | N/A | 0 | 0 | N/A |
| | **bio3** | **2** | **172** | 70 | 70 | 100% | 1 | 1 | 100% | 0 | 0 | N/A |
| | **bio4** | **2** | **170** | 68 | 68 | 100% | 1 | 1 | 100% | 0 | 0 | N/A |
| | **bio5** | **3** | **153** | 62 | 61 | 98.4% | 0 | 0 | N/A | 0 | 0 | N/A |
| | **bio6** | **2** | **167** | 67 | 67 | 100% | 1 | 1 | 100% | 1 | 1 | 100% |
| | **bio7** | **2** | **174** | 65 | 64 | 98.5% | 1 | 1 | 100% | 0 | 0 | N/A |
| | **bio8** | **2** | **174** | 71 | 71 | 100% | 0 | 0 | N/A | 0 | 0 | N/A |
| | **bio9** | **2** | **170** | 67 | 64 | 95.5% | 1 | 1 | 100% | 0 | 0 | N/A |
| | **bio10** | **4** | **166** | 58 | 58 | 100% | 1 | 1 | 100% | 0 | 0 | N/A |
| | **bio11** | **4** | **174** | 73 | 68 | 93.2% | 1 | 1 | 100% | 0 | 0 | N/A |
| | **bio12** | **2** | **174** | 70 | 70 | 100% | 0 | 0 | N/A | 0 | 0 | N/A |
| | **bio13** | **2** | **127** | 47 | 46 | 97.9% | 0 | 0 | N/A | 0 | 0 | N/A |
| | **bio14** | **2** | **172** | 74 | 74 | 100% | 0 | 0 | N/A | 0 | 0 | N/A |
| | **bio15** | **2** | **175** | 73 | 73 | 100% | 0 | 0 | N/A | 0 | 0 | N/A |
| | **bio16** | **3** | **174** | 68 | 67 | 98.5% | 1 | 1 | 100% | 1 | 1 | 100% |
| | **bio17** | **2** | **172** | 71 | 71 | 100% | 1 | 1 | 100% | 1 | 0 | 0% |
| | **bio18** | **2** | **174** | 73 | 72 | 98.6% | 0 | 0 | N/A | 0 | 0 | N/A |
| | **bio19** | **2** | **174** | 78 | 76 | 97.4% | 0 | 0 | N/A | 1 | 0 | 0% |
| | **bio20** | **2** | **174** | 67 | 67 | 100% | 1 | 1 | 100% | 0 | 0 | N/A |
| | **bio21** | **2** | **174** | 68 | 68 | 100% | 1 | 1 | 100% | 1 | 1 | 100% |
| | **bio22** | **2** | **174** | 73 | 73 | 100% | 1 | 1 | 100% | 0 | 0 | N/A |
| | **bio23** | **2** | **163** | 67 | 67 | 100% | 0 | 0 | N/A | 0 | 0 | N/A |
| | **Median** | | | | | **100%** | | | **100%** | | | **50%*** |
| Technical (same specimen, different sample) | **tech1** | **2** | **175** | 69 | 69 | 100% | 1 | 1 | 100% | 0 | 0 | N/A |
| | **tech2** | **2** | **174** | 69 | 69 | 100% | 0 | 0 | N/A | 0 | 0 | N/A |
| | **tech3** | **2** | **174** | 69 | 69 | 100% | 1 | 1 | 100% | 0 | 0 | N/A |
| | **tech4** | **2** | **174** | 64 | 64 | 100% | 1 | 1 | 100% | 0 | 0 | N/A |
| | **Median** | | | | | **100%** | | | **100%** | | | **N/A** |

**Table S1. Mutation-level concordance of biological and technical replicates.** 23 biological replicates (top section: same patient, different specimen) and four technical replicates (bottom section: same specimen, different sample) were evaluated for concordance across three mutation types (substitutions, deletions, and insertions) with median percent concordance shown at the bottom of each section for each mutation type. Each row corresponds to a replicate set of 2–4 samples (# samples) with mutation concordance constrained to annotated, completely unambiguous (i.e., A/T/C/G base calls only) genes (# genes). Each mutation group shows the total number of mutations along with the number and percentage of concordant mutation calls. In sets with >2 samples, if just one sample had a different mutation call, then it was considered discordant. Concordance is shown as N/A when zero mutations were assessed. *Three replicate sets have one insertion called by all samples, while three have one insertion called by some but not all samples. Incidentally, this insertion lies within a tandem repeat: NC_063383.1:136,513-136,569 (ATC).

| Sample ID | # Genes | Substitutions | | | Deletions | | | Insertions | | |
|---|---|---|---|---|---|---|---|---|---|---|
| | | Total | Concordant | | Total | Concordant | | Total | Concordant | |
| | | | # | % | | # | % | | # | % |
| **LC0000050** | 157 | 56 | 56 | 100% | 0 | 0 | N/A | 0 | 0 | N/A |
| **LC0000024** | 159 | 70 | 70 | 100% | 1 | 1 | 100% | 1 | 1 | 100% |
| **LC0000302** | 138 | 48 | 48 | 100% | 0 | 0 | N/A | 1 | 1 | 100% |
| **LC0000056** | 174 | 71 | 71 | 100% | 0 | 0 | N/A | 0 | 0 | N/A |
| **LC0000280** | 173 | 69 | 69 | 100% | 0 | 0 | N/A | 0 | 0 | N/A |
| **LC0000203** | 171 | 69 | 69 | 100% | 1 | 1 | 100% | 0 | 0 | N/A |
| **LC0000027** | 152 | 56 | 56 | 100% | 0 | 0 | N/A | 1 | 0 | 0% |
| **LC0000128** | 156 | 55 | 55 | 100% | 0 | 0 | N/A | 1 | 1 | 100% |
| **LC0000085** | 171 | 66 | 66 | 100% | 1 | 1 | 100% | 0 | 0 | N/A |
| **LC0000016** | 152 | 45 | 45 | 100% | 0 | 0 | N/A | 0 | 0 | N/A |
| **LC0000251** | 172 | 63 | 63 | 100% | 1 | 1 | 100% | 0 | 0 | N/A |
| **LC0000097** | 172 | 72 | 72 | 100% | 1 | 1 | 100% | 1 | 1 | 100% |
| **LC0000291** | 136 | 36 | 36 | 100% | 1 | 1 | 100% | 0 | 0 | N/A |
| **LC0000147** | 174 | 78 | 78 | 100% | 0 | 0 | N/A | 0 | 0 | N/A |
| **LC0000019** | 174 | 77 | 77 | 100% | 0 | 0 | N/A | 0 | 0 | N/A |
| **LC0000191** | 174 | 70 | 70 | 100% | 0 | 0 | N/A | 0 | 0 | N/A |
| **LC0000059** | 174 | 68 | 68 | 100% | 1 | 1 | 100% | 1 | 1 | 100% |
| **LC0000305** | 174 | 67 | 67 | 100% | 1 | 1 | 100% | 1 | 1 | 100% |
| **Median** | | **1,136** | **1,136** | **100%** | **8** | **8** | **100%** | **7** | **6** | **85.7%** |

**Table S2. Mutation-level concordance of Labcorp and CDC sequence MPXV genomes.** 18 parallelly sequenced MPXV genomes were evaluated for concordance across three mutation types (substitutions, deletions, and insertions) with median percent concordance shown at the bottom for each mutation type. Each row corresponds to a genome comparison with mutation concordance constrained to annotated, completely unambiguous (i.e., A/T/C/G base calls only) genes (# genes). Each mutation group shows the total number of mutations along with the number and percentage of concordant mutation calls. Concordance is shown as N/A when zero mutations were assessed.

| Clade IIb Lineage | # Sequences | 5X | | 10X | | 20X | | 30X | |
|---|---|---|---|---|---|---|---|---|---|
| | | # Correct | Acc. [%] | # Correct | Acc. [%] | # Correct | Acc. [%] | # Correct | Acc. [%] |
| **A** | 6 | 6 | 100% | 6 | 100% | 6 | 100% | 6 | 100% |
| **A.1** | 6 | 6 | 100% | 6 | 100% | 6 | 100% | 6 | 100% |
| **A.2** | 6 | 6 | 100% | 6 | 100% | 6 | 100% | 6 | 100% |
| **A.2.1** | 6 | 6 | 100% | 6 | 100% | 6 | 100% | 6 | 100% |
| **A.2.2** | 6 | 6 | 100% | 6 | 100% | 6 | 100% | 6 | 100% |
| **A.2.3** | 6 | 6 | 100% | 6 | 100% | 6 | 100% | 6 | 100% |
| **B.1** | 6 | 6 | 100% | 6 | 100% | 6 | 100% | 6 | 100% |
| **B.1.1** | 6 | 5 | ***83.3%*** | 6 | 100% | 6 | 100% | 6 | 100% |
| **B.1.10** | 6 | 6 | 100% | 6 | 100% | 6 | 100% | 6 | 100% |
| **B.1.11** | 6 | 4 | ***66.7%*** | 6 | 100% | 6 | 100% | 6 | 100% |
| **B.1.12** | 6 | 6 | 100% | 6 | 100% | 6 | 100% | 6 | 100% |
| **B.1.13** | 3 | 3 | 100% | 3 | 100% | 3 | 100% | 3 | 100% |
| **B.1.14** | 6 | 2 | ***33.3%*** | 6 | 100% | 6 | 100% | 6 | 100% |
| **B.1.15** | 3 | 2 | ***66.7%*** | 3 | 100% | 3 | 100% | 3 | 100% |
| **B.1.16** | 6 | 6 | 100% | 6 | 100% | 6 | 100% | 6 | 100% |
| **B.1.19** | 6 | 6 | 100% | 6 | 100% | 6 | 100% | 6 | 100% |
| **B.1.2** | 6 | 6 | 100% | 6 | 100% | 6 | 100% | 6 | 100% |
| **B.1.20** | 6 | 6 | 100% | 6 | 100% | 6 | 100% | 6 | 100% |
| **B.1.22** | 3 | 3 | 100% | 3 | 100% | 3 | 100% | 3 | 100% |
| **B.1.3** | 6 | 6 | 100% | 6 | 100% | 6 | 100% | 6 | 100% |
| **B.1.4** | 6 | 6 | 100% | 6 | 100% | 6 | 100% | 6 | 100% |
| **B.1.5** | 6 | 6 | 100% | 6 | 100% | 6 | 100% | 6 | 100% |
| **B.1.6** | 6 | 4 | ***66.7%*** | 6 | 100% | 6 | 100% | 6 | 100% |
| **B.1.7** | 6 | 6 | 100% | 6 | 100% | 6 | 100% | 6 | 100% |
| **B.1.8** | 6 | 6 | 100% | 6 | 100% | 6 | 100% | 6 | 100% |
| **B.1.9** | 6 | 4 | ***66.7%*** | 6 | 100% | 6 | 100% | 6 | 100% |
| **C.1** | 6 | 6 | 100% | 6 | 100% | 6 | 100% | 6 | 100% |
| **C.1.1** | 6 | 5 | ***83.3%*** | 6 | 100% | 6 | 100% | 6 | 100% |
| **E.1** | 6 | 6 | 100% | 6 | 100% | 6 | 100% | 6 | 100% |
| **E.2** | 6 | 6 | 100% | 6 | 100% | 6 | 100% | 6 | 100% |
| **E.3** | 6 | 6 | 100% | 6 | 100% | 6 | 100% | 6 | 100% |
| **F.1** | 6 | 6 | 100% | 6 | 100% | 6 | 100% | 6 | 100% |
| **F.2** | 6 | 6 | 100% | 6 | 100% | 6 | 100% | 6 | 100% |
| **F.3** | 3 | 3 | 100% | 3 | 100% | 3 | 100% | 3 | 100% |
| **F.4** | 3 | 3 | 100% | 3 | 100% | 3 | 100% | 3 | 100% |
| **Total** | **195** | **182** | **93%** | **195** | **100%** | **195** | **100%** | **195** | **100%** |

**Table S3. Clade IIb simulation lineage accuracy results.** Each row shows the number (#) of sequences simulated for each lineage along with the number (#) correct and % accuracy (acc.) for each depth of coverage simulated (5X, 10X, 20X, 30X).
